# Intended Isocaloric Time-Restricted Eating Shifts Circadian Clocks but Does Not Improve Cardiometabolic Health in Women with Overweight

**DOI:** 10.1101/2024.10.05.24314120

**Authors:** Beeke Peters, Julia Schwarz, Bettina Schuppelius, Agnieszka Ottawa, Daniela A. Koppold, Daniela Weber, Nico Steckhan, Knut Mai, Tilman Grune, Andreas F.H. Pfeiffer, Andreas Michalsen, Achim Kramer, Olga Pivovarova-Ramich

## Abstract

Time-restricted eating (TRE) is a promising strategy to improve metabolic outcomes. However, it remains unclear whether TRE has cardiometabolic benefits in an isocaloric setting and whether its effects depend on the eating timing. We conducted a randomized cross-over trial to directly compare the effects of a two-week early TRE (eTRE: eating between 8:00 and 16:00 hr) and a two-week late TRE (lTRE: eating between 13:00 and 21:00 hr) on insulin sensitivity, cardiometabolic risk factors, and the internal circadian phase. During the restricted 8-hour eating period, participants were asked to consume their habitual food quality and quantity. In 31 women with overweight or obesity, insulin sensitivity did not differ between (−0.07; 95% CI, −0.77 to 0.62, P = 0.60) and within (eTRE: 0.31; 95% CI, −0.14 to 0.76, P = 0.11; lTRE: 0.19; 95% CI, −0.22 to 0.60, P = 0.25) interventions. 24-hour glucose, lipid, inflammatory, and oxidative stress markers showed no clinically meaningful between- and within-intervention differences. Participants demonstrated high timely adherence (eTRE: 96.5%, lTRE: 97.7%), unchanged dietary composition and physical activity, minor daily calorie deficit (eTRE: −167 kcal/day) and weight loss (eTRE: −1.08 kg, lTRE: −0.44 kg). lTRE delayed the circadian phase in blood monocytes (24 min; 95% CI, −5 to 54 min, *P* = 0.10) and sleep midpoint (15 min; 95% CI, 7 to 22 min, P = 0.001) compared to eTRE. Overall, in an intended isocaloric setting, neither eTRE nor lTRE improve insulin sensitivity or other cardiometabolic traits despite a shift of internal circadian clocks.

**One sentence summary:** Nearly isocaloric time-restricted eating shifts circadian clocks but does not improve cardiometabolic health in women with overweight or obesity

## Introduction

Time-restricted eating (TRE) is a form of intermittent fasting characterized by a daily eating window of 10 hours or less and a prolonged fasting period of at least 14 hours over the course of the day (*1*). TRE is becoming increasingly popular as a simple dietary approach to control body weight and improve metabolic health (*2, 3*). In rodents, TRE is protective against diet-induced obesity and associated metabolic disturbances (*4, 5*). Similarly, human trials on TRE have highlighted numerous beneficial cardiometabolic effects, such as improved fasting (*3, 6, 7*) and mean daily (*3, 8*) glucose concentrations, insulin resistance (*3, 7, 9*) or insulin sensitivity (*2, 10*), triglyceride (*6, 8, 11, 12*), total (*12, 13*) and low-density-lipoprotein (LDL) cholesterol (*13*) concentrations, and blood pressure (*2, 13, 14*) as well as moderate body weight (*8, 13, 15–17*) and body fat reduction (*7, 13, 17*). Therefore, beyond its effects on body weight, TRE represents a promising approach to combat insulin resistance and diabetes.

However, results of TRE trials are inconsistent (*18, 19*) and require stronger clinical trial evidence (*20*) to answer several practically relevant questions. Especially, it is unclear whether metabolic improvements are induced by the restriction of the daily eating duration itself, by accompanying caloric restriction (and respective weight loss), or by the combination of both factors. Indeed, most TRE trials did not carefully monitor energy intake and/or other potential cofounders. Therefore, we investigated in a clinical trial whether 8-hour TRE can improve insulin sensitivity and other cardiometabolic parameters in an intended isocaloric setting, where we precisely controlled calorie intake, timely adherence, dietary composition, and physical activity.

The secondary objective of the trial was to compare the effects of eating early (eTRE) vs. late (lTRE) in the day during the TRE intervention. Although most clinical studies suggest additional benefits of eTRE (*2, 3, 10, 21*), which might be explained by circadian rhythms of key metabolic processes (*19, 22*), trials that directly compare eTRE and lTRE are very limited (*8, 23, 24*). Based on previous research, we hypothesized that an isocaloric TRE would improve insulin sensitivity and cardiometabolic health compared to baseline and that eTRE would be more effective than lTRE. To investigate this, we conducted a 10-week randomized crossover ChronoFast trial (NCT04351672) including two 2-week isocaloric dietary intervention periods: (1) eTRE (8-hour eating window between 8:00 and 16:00 hr) and (2) lTRE (8-hour eating window between 13:00 and 21:00 hr), preceded by a 4-week baseline period and separated by a 2-week washout period (*18*) (**Fig. 1A**). Because some previously published trials, comparing eTRE and lTRE or investigating impacts of TRE on glucose metabolism, have included exclusively male subjects (*2, 7, 8, 10*) and therefore may not be generalizable to females, our trial was conducted exclusively in women with overweight or obesity. Here, we report the trial results on insulin sensitivity and other cardiometabolic outcomes as well as internal circadian time.

**Fig. 1.**
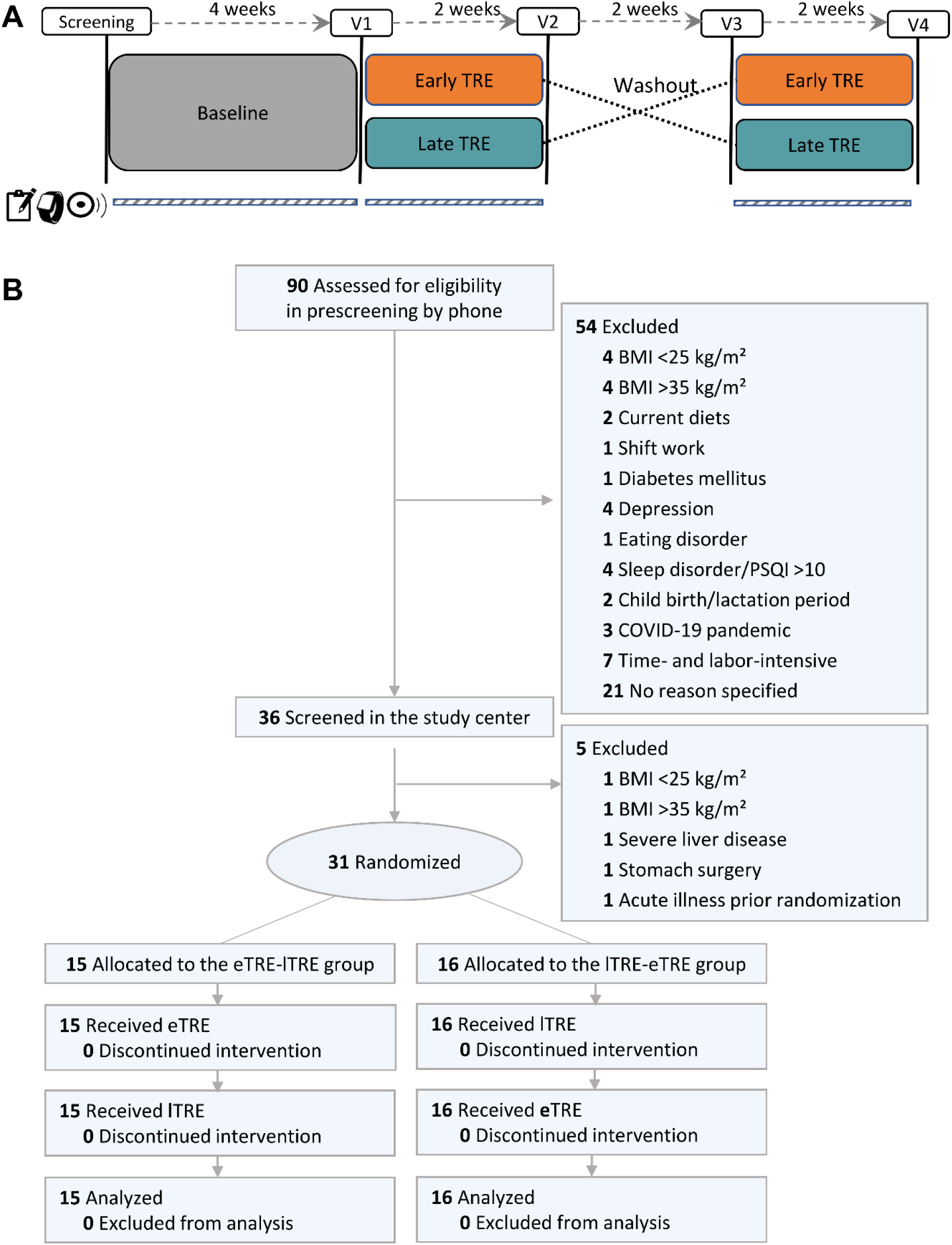
Study Design and Participant Flow Diagram. A, study design. The ChronoFast study was a 10-week randomized crossover trial including two 2-week dietary intervention periods: (1) early time-restricted eating (eTRE: 8-hour eating window between 8:00 and 16:00 hr) and (2) late time-restricted eating (lTRE: 8-hour eating window between 13:00 and 21:00 hr), preceded by a 4-week baseline period and separated by a 2-week washout period. Prior to baseline, participants were pre-screened by phone and completed comprehensive screening at the study center to determine eligibility. Eligible participants were randomly allocated to the eTRE-lTRE or lTRE-eTRE study arms based on their BMI and age. During the study center visits (V1-4) before and after each intervention, glycemic and other cardiometabolic parameters were assessed in a fasting state and in an oral glucose tolerance test. During the 14 days of the baseline and both TRE intervention periods, continuous glucose monitoring (CGM), actigraphy, food and sleep diaries were conducted. B, CONSORT diagram describing a number of participants throughout the study, from enrollment to completion.

## Results

### Participants Characteristics and Study Flow

Between April 2020 to December 2021, we screened 90 and enrolled 31 participants, of whom 15 were allocated to the eTRE-lTRE and 16 to the lTRE-eTRE study arms (**Fig. 1B**). Participants had a mean (Standard Deviation, SD), Body Mass Index (BMI) of 30.5 (2.9) and median (Interquartile Range, IQR) age of 62 (53–65) years. 18 participants showed a normal (NGT), and 13 - an impaired fasting glucose or impaired glucose tolerance (IFG/IGT). All participants were female (26 postmenopausal and 5 pre-menopausal), White, and of Caucasian ethnicity (**Table 1**). Despite the COVID-19 pandemic, there were no dropouts after randomization, and all 31 participants completed the study. Final analysis included 31 subjects.

**Table 1.**
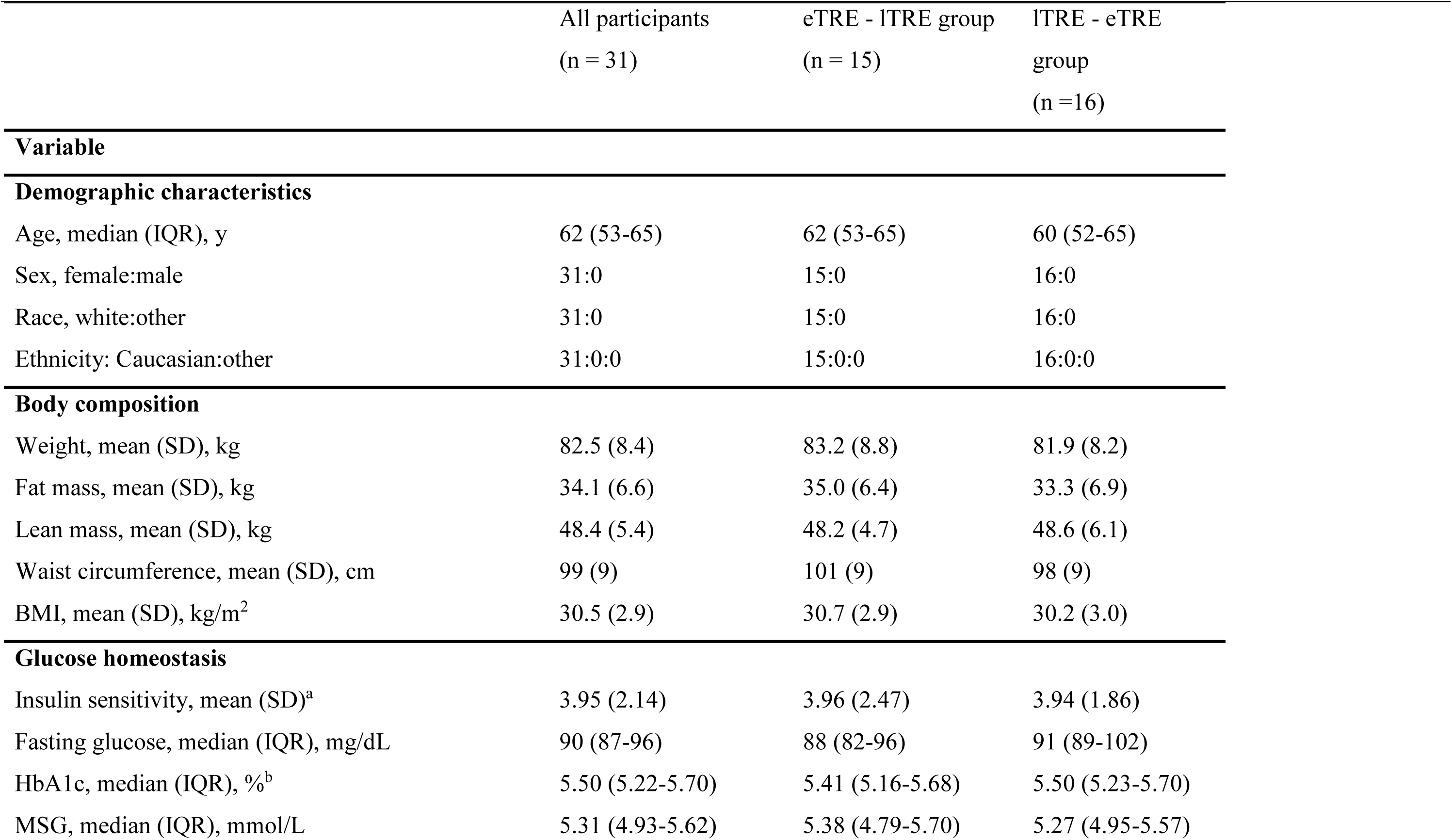
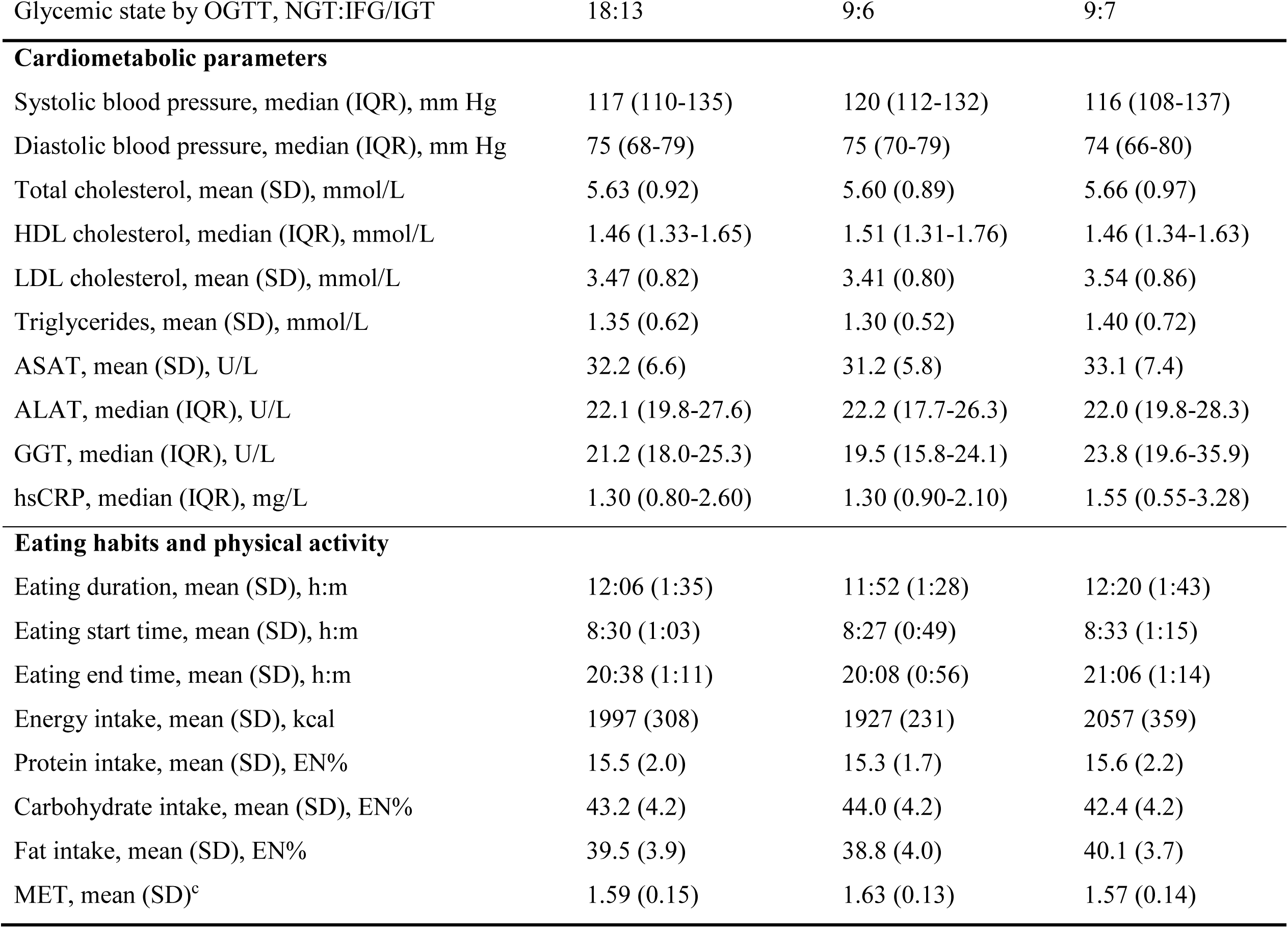
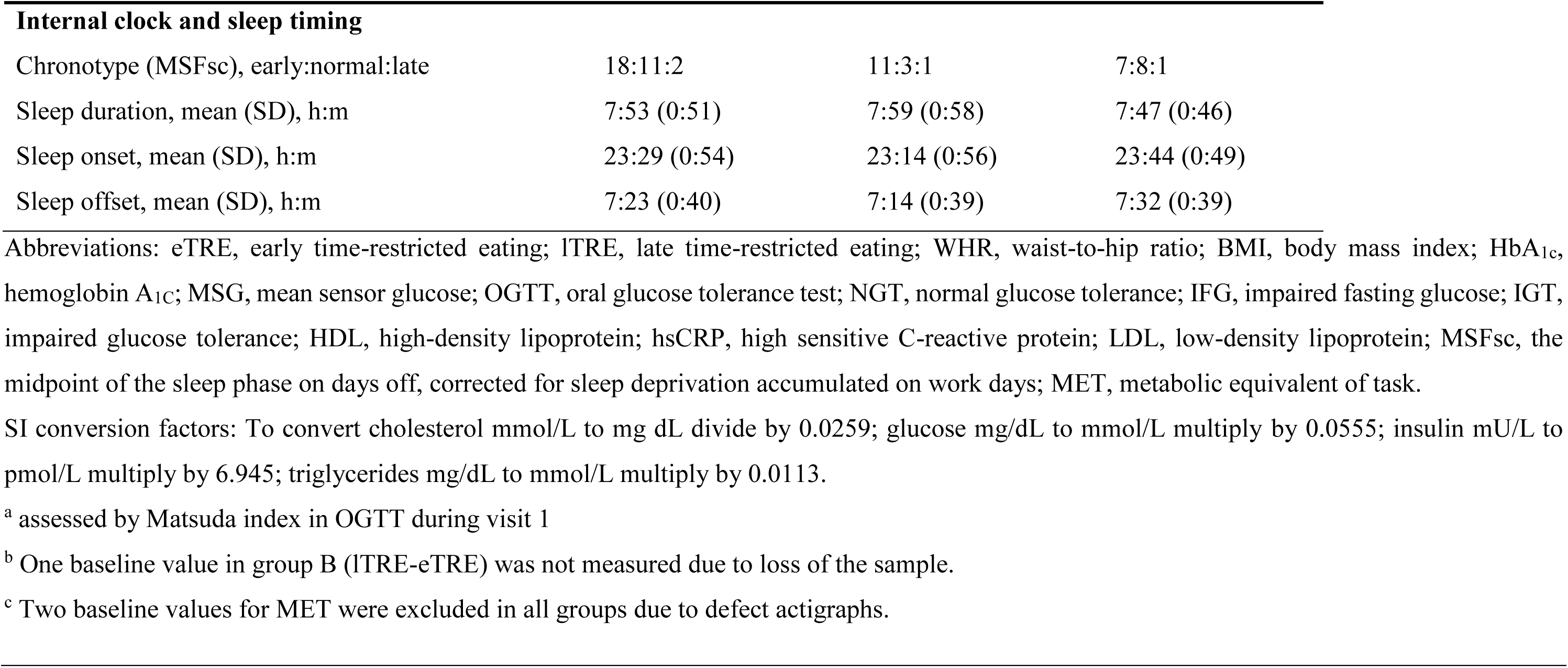
Baseline Characteristics of Study Participants.

|  | All participants<br>(n = 31) | eTRE - lTRE group<br>(n = 15) | lTRE - eTRE<br>group<br>(n =16) |
| --- | --- | --- | --- |
| **Variable** |  |  |  |
| **Demographic characteristics** |  |  |  |
| Age, median (IQR), y | 62 (53-65) | 62 (53-65) | 60 (52-65) |
| Sex, female:male | 31:0 | 15:0 | 16:0 |
| Race, white:other | 31:0 | 15:0 | 16:0 |
| Ethnicity: Caucasian:other | 31:0:0 | 15:0:0 | 16:0:0 |
| **Body composition** |  |  |  |
| Weight, mean (SD), kg | 82.5 (8.4) | 83.2 (8.8) | 81.9 (8.2) |
| Fat mass, mean (SD), kg | 34.1 (6.6) | 35.0 (6.4) | 33.3 (6.9) |
| Lean mass, mean (SD), kg | 48.4 (5.4) | 48.2 (4.7) | 48.6 (6.1) |
| Waist circumference, mean (SD), cm | 99 (9) | 101 (9) | 98 (9) |
| BMI, mean (SD), kg/m<sup>2</sup> | 30.5 (2.9) | 30.7 (2.9) | 30.2 (3.0) |
| **Glucose homeostasis** |  |  |  |
| Insulin sensitivity, mean (SD)<sup>a</sup> | 3.95 (2.14) | 3.96 (2.47) | 3.94 (1.86) |
| Fasting glucose, median (IQR), mg/dL | 90 (87-96) | 88 (82-96) | 91 (89-102) |
| HbA1c, median (IQR), %<sup>b</sup> | 5.50 (5.22-5.70) | 5.41 (5.16-5.68) | 5.50 (5.23-5.70) |
| MSG, median (IQR), mmol/L | 5.31 (4.93-5.62) | 5.38 (4.79-5.70) | 5.27 (4.95-5.57) |
| Glycemic state by OGTT, NGT:IFG/IGT | 18:13 | 9:6 | 9:7 |
| **Cardiometabolic parameters** |  |  |  |
| Systolic blood pressure, median (IQR), mm Hg | 117 (110-135) | 120 (112-132) | 116 (108-137) |
| Diastolic blood pressure, median (IQR), mm Hg | 75 (68-79) | 75 (70-79) | 74 (66-80) |
| Total cholesterol, mean (SD), mmol/L | 5.63 (0.92) | 5.60 (0.89) | 5.66 (0.97) |
| HDL cholesterol, median (IQR), mmol/L | 1.46 (1.33-1.65) | 1.51 (1.31-1.76) | 1.46 (1.34-1.63) |
| LDL cholesterol, mean (SD), mmol/L | 3.47 (0.82) | 3.41 (0.80) | 3.54 (0.86) |
| Triglycerides, mean (SD), mmol/L | 1.35 (0.62) | 1.30 (0.52) | 1.40 (0.72) |
| ASAT, mean (SD), U/L | 32.2 (6.6) | 31.2 (5.8) | 33.1 (7.4) |
| ALAT, median (IQR), U/L | 22.1 (19.8-27.6) | 22.2 (17.7-26.3) | 22.0 (19.8-28.3) |
| GGT, median (IQR), U/L | 21.2 (18.0-25.3) | 19.5 (15.8-24.1) | 23.8 (19.6-35.9) |
| hsCRP, median (IQR), mg/L | 1.30 (0.80-2.60) | 1.30 (0.90-2.10) | 1.55 (0.55-3.28) |
| **Eating habits and physical activity** |  |  |  |
| Eating duration, mean (SD), h:m | 12:06 (1:35) | 11:52 (1:28) | 12:20 (1:43) |
| Eating start time, mean (SD), h:m | 8:30 (1:03) | 8:27 (0:49) | 8:33 (1:15) |
| Eating end time, mean (SD), h:m | 20:38 (1:11) | 20:08 (0:56) | 21:06 (1:14) |
| Energy intake, mean (SD), kcal | 1997 (308) | 1927 (231) | 2057 (359) |
| Protein intake, mean (SD), EN% | 15.5 (2.0) | 15.3 (1.7) | 15.6 (2.2) |
| Carbohydrate intake, mean (SD), EN% | 43.2 (4.2) | 44.0 (4.2) | 42.4 (4.2) |
| Fat intake, mean (SD), EN% | 39.5 (3.9) | 38.8 (4.0) | 40.1 (3.7) |
| MET, mean (SD)<sup>c</sup> | 1.59 (0.15) | 1.63 (0.13) | 1.57 (0.14) |
| **Internal clock and sleep timing** |  |  |  |
| Chronotype (MSFsc), early:normal:late | 18:11:2 | 11:3:1 | 7:8:1 |
| Sleep duration, mean (SD), h:m | 7:53 (0:51) | 7:59 (0:58) | 7:47 (0:46) |
| Sleep onset, mean (SD), h:m | 23:29 (0:54) | 23:14 (0:56) | 23:44 (0:49) |
| Sleep offset, mean (SD), h:m | 7:23 (0:40) | 7:14 (0:39) | 7:32 (0:39) |

Abbreviations: eTRE, early time-restricted eating; lTRE, late time-restricted eating; WHR, waist-to-hip ratio; BMI, body mass index; HbA<sub>1c</sub>, hemoglobin A<sub>1c</sub>; MSG, mean sensor glucose; OGTT, oral glucose tolerance test; NGT, normal glucose tolerance; IFG, impaired fasting glucose; IGT, impaired glucose tolerance; HDL, high-density lipoprotein; hsCRP, high sensitive C-reactive protein; LDL, low-density lipoprotein; MSFsc, the midpoint of the sleep phase on days off, corrected for sleep deprivation accumulated on work days; MET, metabolic equivalent of task.
SI conversion factors: To convert cholesterol mmol/L to mg dL divide by 0.0259; glucose mg/dL to mmol/L multiply by 0.0555; insulin mU/L to pmol/L multiply by 6.945; triglycerides mg/dL to mmol/L multiply by 0.0113.
<sup>a</sup> assessed by Matsuda index in OGTT during visit 1
<sup>b</sup> One baseline value in group B (lTRE-eTRE) was not measured due to loss of the sample.
<sup>c</sup> Two baseline values for MET were excluded in all groups due to defect actigraphs.

30 adverse events possibly related to the study were reported, but none were serious (**Table S1**).

### Adherence to TRE Interventions and Study Protocol

All participants showed high adherence to the 8-hour eating duration during the TRE interventions as shown by analysis of food records over 14 days during the baseline and both interventions. During the baseline, they ate within a mean (SD) time period of 12:06 (1:35) hours. The eating duration was mean (SD) of 7:09 (0:32) hours in eTRE and of 6:57 (0:50) hours in lTRE (**Fig. 2A, Fig. S1**). Adherence to the prescribed eating time was also high, with a mean (SD) of 96.5% (6.3%) in eTRE and 97.7% (6.1%) intervention days in lTRE, according to the food record analysis (**Fig. 2B**). Energy intake remained unchanged in lTRE (−97 kcal; 95% Confidence Interval (CI), −193 to −2; P = 0.06) but decreased minimally in eTRE (−167 kcal; 95% CI, −249 to −86 kcal; *P* < 0.001) compared to the baseline (**Fig. 2C)**. No changes in percentage of carbohydrate, fat, and protein intake, as well as in physical activity levels, were found in both TRE interventions compared to the baseline (**Fig. 2D-E, Table S2**).

**Fig. 2.**
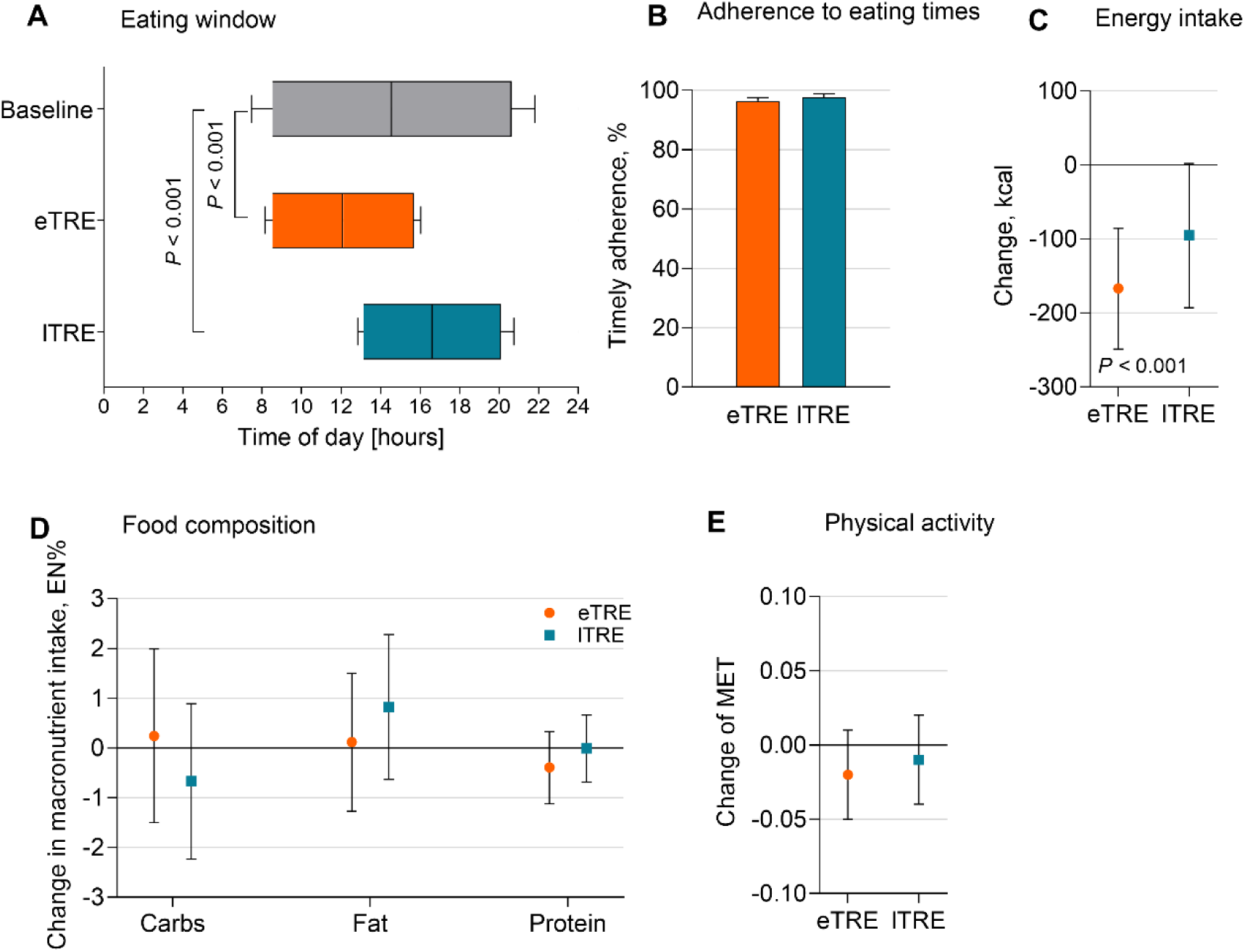
Adherence to eTRE and lTRE Regimes. A, Shown are the times of day (mean [SD]) when participants began eating (left end of box and left whisker) and stopped eating (right end of box and right whisker) in the baseline phase, eTRE and lTRE intervention phases. The vertical line within the boxes indicates the midpoint of the eating window (averaged across all participants). B, Timely adherence defined as a percentage of the adherent timing of eTRE and lTRE to specified times (eTRE: 8:00 to 16:00 ± 30 min and lTRE: 13:00 to 21:00 ± 30 min). Data are shown as means [SD] (bars and whiskers). C, Displayed are changes in energy intake compared to the baseline. Data are shown as means with 95% CI. D, Food composition defined as macronutrient intake. Data presenting changes within eTRE and lTRE are shown as means with 95% CI. E, Physical activity defined as MET. Data presenting changes are shown as means with 95% CI.

### Body Weight and Composition

Participants showed a minimal weight loss of −1.08 kg (95% CI, −0.77 to −1.40 kg; *P* <0.001) within eTRE and −0.44 kg (95% CI, −0.74 to −0.13; *P* = 0.01) within lTRE resulting in a between-intervention difference of 0.65 kg (95% CI, 0.27 to 1.03 kg; *P* = 0.012) (**Table 2**). BMI decreased in eTRE (−0.45 kg/m^2^; 95% CI, −0.56 to −0.33 kg/m^2^; *P* <0.001) and lTRE (−0.12 kg/m^2^; 95% CI, −0.21 to −0.03 kg/m^2^; *P* = 0.01) with a between-diet difference of 0.33 kg/m^2^ (95% CI, 0.21 to 0.44 kg/m^2^; *P* < 0.001). Fat mass loss (−0.61 kg, 95% CI, −1.01 to −0.22 kg; P= 0.002) and lean mass loss (−0.57 kg; −1.11 to −0.04 kg; *P* = 0.04) were observed within eTRE only, but their percentage to body weight was not changed within and between both interventions (**Table 2**).

**Table 2.**
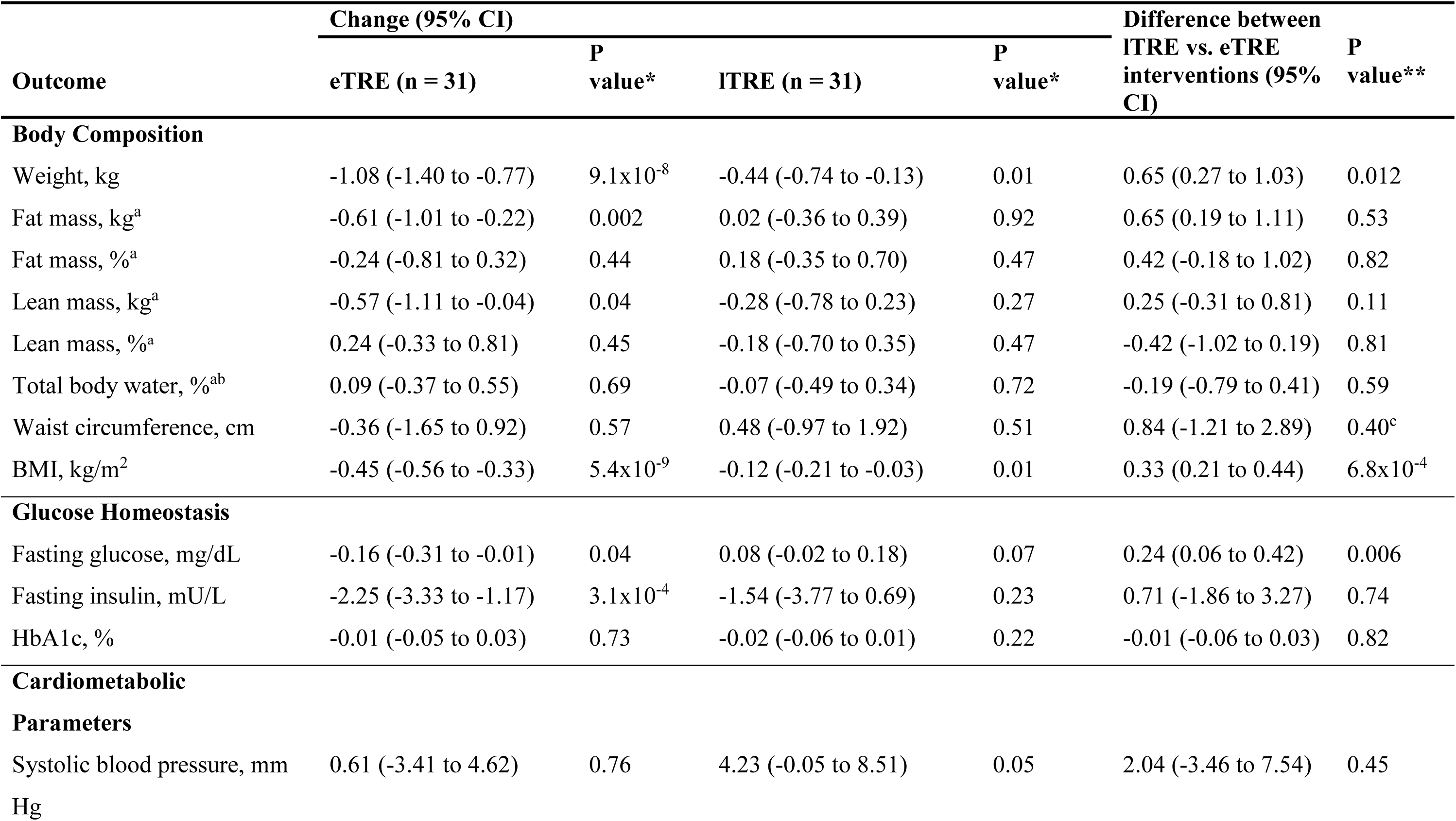
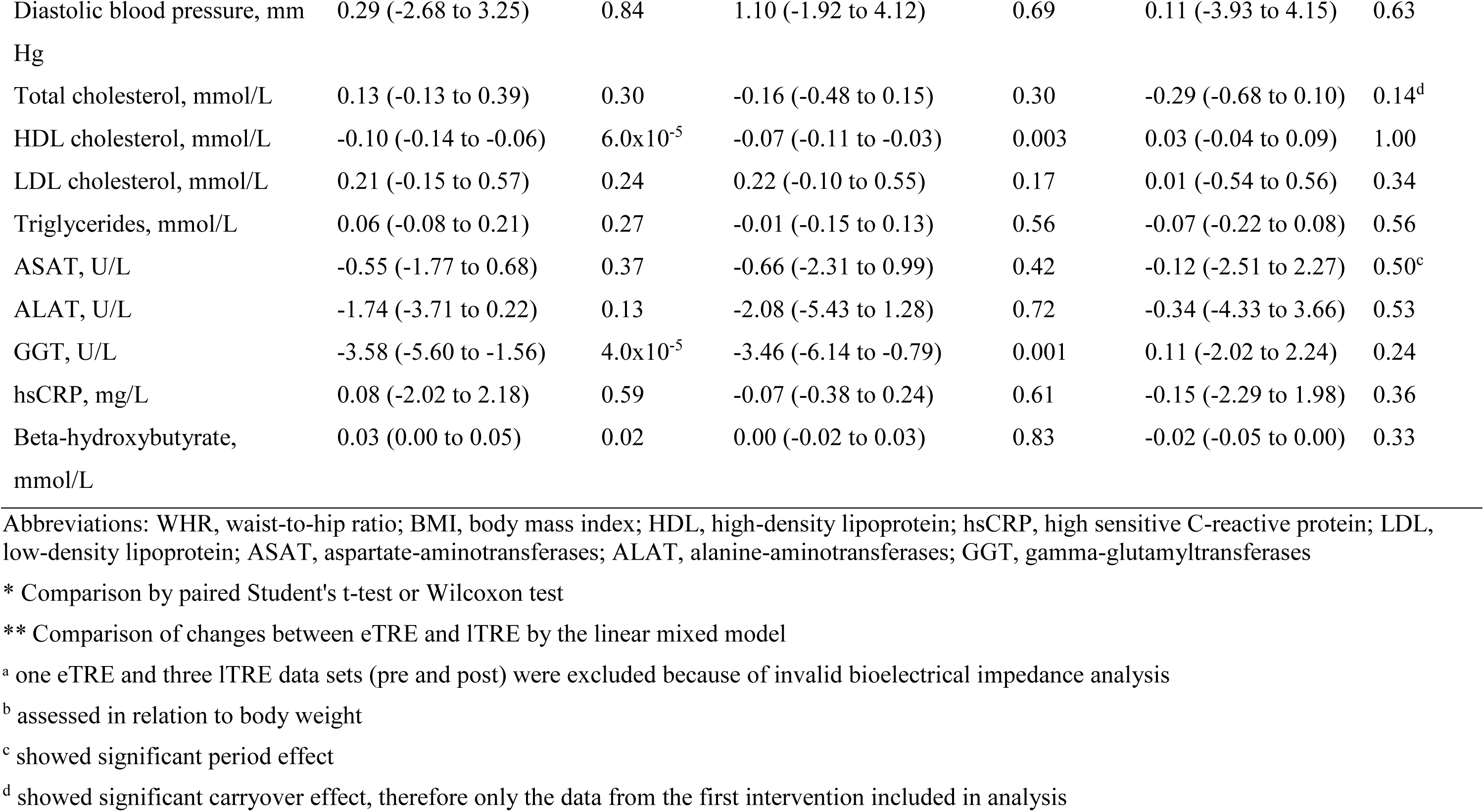
TRE Effects on Body Composition, Glucose Homeostasis, and Cardiometabolic Parameters.

### Insulin Sensitivity and Glucose Homeostasis

Insulin sensitivity assessed by Matsuda index showed no differences between the TRE interventions (−0.07; 95% CI, −0.77 to 0.62, *P* = 0.60 for lTRE vs. eTRE) as well as within eTRE (0.31; 95% CI, −0.14 to 0.76, *P* = 0.11) and lTRE (0.19; 95% CI, −0.22 to 0.60, *P* = 0.25) (**Fig. 3A**). Similarly, the post hoc analysis of the oral glucose insulin sensitivity (OGIS) index revealed no changes within (eTRE: −0.35; 95% CI, −15.2 to 14.5; *P* = 0.54; lTRE: −2.68; 95% CI, −15.9 to 10.6; *P* = 0.80) and between-interventions (−2.32; 95% CI, −23.3 to 18.7; *P* = 0.67). Area under the curve (AUC) glucose in oral glucose tolerance test (OGTT) increased within eTRE (1636; 95% CI, 798 to 2475, *P* = 0.001), and its changes differed between interventions (−2175; 95% CI, −3088 to 1262, *P* = 0.002) (**Fig. S2A-B**). This minor decline in OGTT-derived glucose tolerance within eTRE might be explained by a decrease of insulin secretion relative to glucose, as indicated by an insulinogenic index (−0.43; 95% CI, −0.69 to −0.16; *P* <0.001; **Fig. S2C-E**). This was also reflected by a decrease in the disposition index (−1.58; 95% CI, −2.75 to −0.41, *P* = 0.01**; Fig. S2F**), by an increase of free fatty acids (**Fig. S2K-L)** and aligns with a decrease of glucagon within eTRE as derived in OGTT (**Fig. S2I-J)**. Despite minor changes in glucose tolerance in OGTT, mean 24-hour glucose in continuous glucose monitoring (CGM) showed no differences between and no changes within eTRE and lTRE compared to baseline (**Fig. 3B-C, Table S3**). Glucose variability in CGM showed increased intra-day variation and prolonged time in the high-glucose range within eTRE, while inter-day variation decreased within lTRE (**Table S3**).

**Fig. 3.**
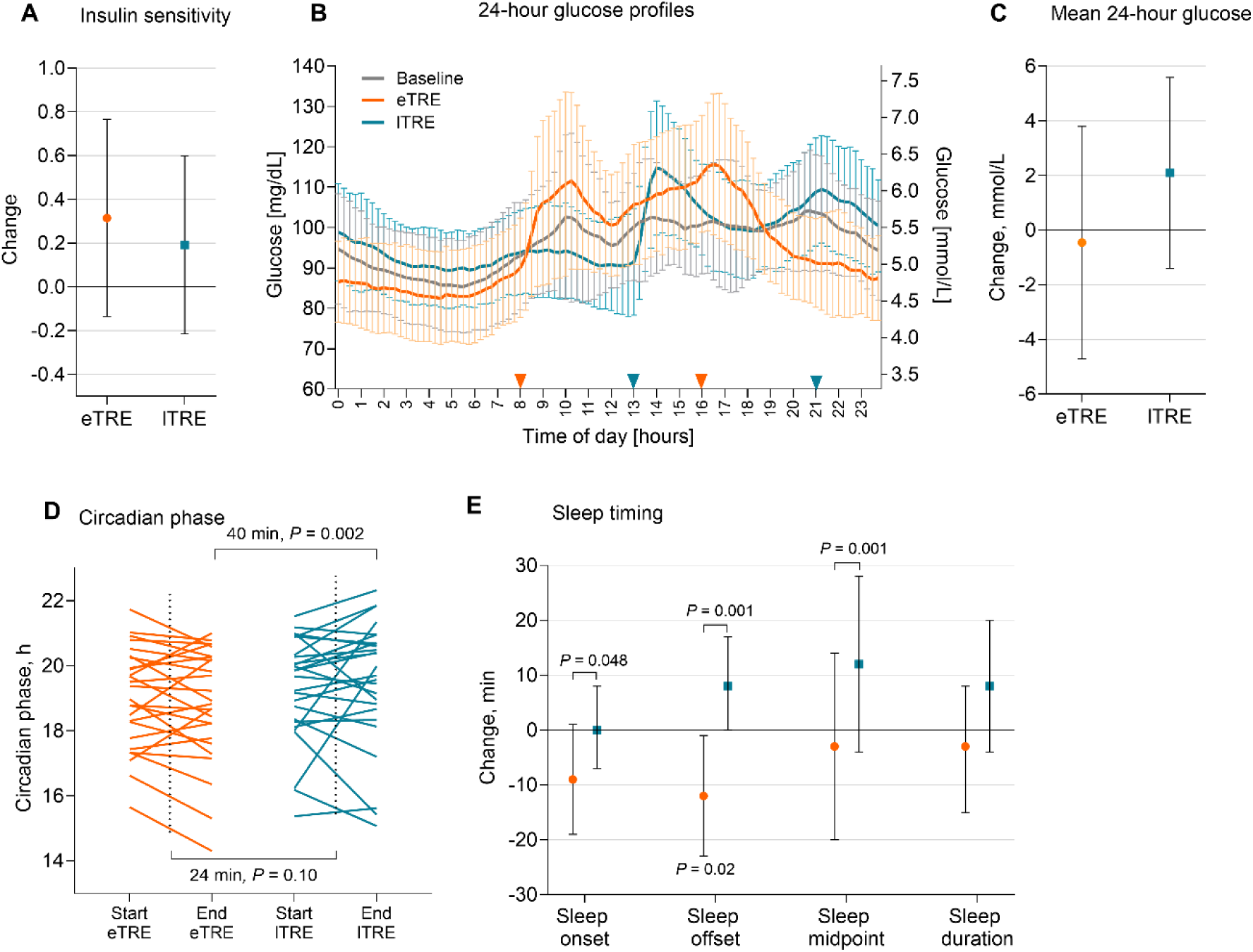
Effects of eTRE and lTRE on Glucose Homeostasis and Circadian Time. A, Plotted are changes in insulin sensitivity in eTRE and lTRE. Data are shown as means with 95% CI. B, Plotted are 24-hour glucose profiles assessed by CGM over 14 days in the baseline phase, eTRE and lTRE intervention phase. Solid lines and whiskers represent the mean glucose and SD, respectively, summarizing all assessed days for 30 participants. C, Plotted are the mean 24-hour glucose values assessed by CGM over 14 days in the baseline phase, eTRE and lTRE intervention phase. Data are shown as means with 95% CI. D, Circadian phase before and after eTRE and lTRE assessed by BodyTime assay in blood monocytes. Dashed black lines show between-intervention changes. Black solid P-value-bar shows comparison of samples collected after both interventions (n=26). Orange and petrol lines bind individual circadian phase values before and after eTRE and lTRE, respectively. The circadian phase corresponds to the BodyTime-predicted DLMO. E, Presented are changes of sleep timing showing that sleep onset, sleep offset, and sleep midpoint within eTRE and lTRE. Data are shown as means with 95% CI.

### Cardiometabolic Parameters

None of the TRE interventions affected systolic and diastolic blood pressure, total cholesterol, low density lipoprotein (LDL) cholesterol, and triglyceride concentrations (**Table 2**). High-density lipoprotein (HDL) cholesterol declined within both eTRE (−0.10 mmol/L; 95% CI, - 0.14 to −0.06 mmol/L; *P* <0.001) and lTRE (−0.07 mmol/L; 95% CI, −0.11 to −0.03 mmol/L, *P* = 0.003), with no difference between the interventions. Both the eTRE (−3.58 U/L; 95% CI, - 5.60 to −1.56 U/L, *P* <0.001) and the lTRE (−3.46 U/L; 95% CI −6.14 to −0.79 U/L, *P* = 0.001) induced a decrease in γ-glutamyltransferase (GGT) but not in aspartate aminotransferase (ALT) and alanine aminotransferase (AST) (**Table 2**).

### Adipokines, Inflammatory Markers, and Oxidative Stress Markers

We further investigated concentrations of adipokines leptin and adiponectin, as well as inflammatory markers interleukin 6 (IL-6), tumor necrosis factor alpha (TNFα), monocyte chemotactic protein 1 (MCP-1), and high-sensitive C-reactive protein (hsCRP), which strongly contribute to obesity and diabetes pathogenesis. Leptin concentration declined within both eTRE (−8080 pg/mL; 95% CI −15995 to −166 pg/mL, *P* = 0.02) and lTRE (−10763 pg/mL; 95% CI −19035 to −2492 pg/mL; *P* = 0.001) without differences between interventions. Adiponectin concentrations were also reduced within eTRE (−0.94 pg/mL; 95% CI −1.54 to 0.34 pg/mL, *P* = 0.003) without differences between interventions (**Fig. S3A-B)**. The inflammatory markers were not affected by either eTRE or lTRE intervention (**Table 2, Fig. S3C-E)**. Concentrations of oxidative stress markers malondialdehyde, 3-nitrotyrosine, and protein carbonyls were not changed in any TRE intervention (**Table S4**).

### Hunger and Satiety

Participants showed lower desire to eat, hunger, and capacity to eat during the lTRE in the morning, but not in the evening compared to the eTRE whereas satiety did not differ at any time of day (**Table S5**). In agreement with this, changes in the satiety hormone peptide YY (PYY) differed between interventions (34.7 pg/mL; 95% CI, 18.2 to 51.3 pg/mL, *P* <0.001), showing a decrease within eTRE (−13.0 pg/mL; 95% CI −24.5 to −1.5 pg/mL, *P* = 0.01) and increase within lTRE (22.5 pg/mL; 95% CI 13.1 to 31.9 pg/mL, P <0.001). Concentrations of the hunger hormone ghrelin remained unchanged in both TRE interventions (**Table S5**).

### Analysis in participants with impaired glucose metabolism

Additional analysis in a subcohort with impaired glucose metabolism (IFG/IFT) revealed no differences in the insulin sensitivity and mean 24-hour glucose between and within TRE interventions. Other secondary glycemic, cardiometabolic, inflammatory, and oxidative stress outcomes also showed effects similar to the whole cohort (**Table S6, Table S7**).

### PBMC Gene Expression

To elucidate molecular pathway potentially induced by TRE, we then measured the expression of genes coding inflammatory markers (*IL6, TNFα, CCL2, IL10*), as well as key metabolic genes (*CPT1A, PDK4, SIRT1, FASN, LPL*), and clock genes (*CLOCK, BMAL1, PER1, PER2, NR1D1, CRY1, CRY2, RORA*) in peripheral blood mononuclear (PBMC) samples (**Table S8**). Core clock genes *PER1* (−0.30; 95% CI, −0.57 to −0.03, *P* = 0.03) and *NR1D1* (−0.24; 95% CI, - 0.48 to −0.01, *P* = 0.02), which are strongly involved in metabolic regulation (*25*), declined their expression within eTRE without between-intervention difference. Other genes showed no TRE-induced expression changes (**Fig. S4**).

### Sleep Timing and Circadian Phase

Considering the data on the effect of food intake on circadian clocks (*24, 26–28*) and changes in the PBMC gene expression, we tested whether the eating timing during the TRE affects internal circadian phase. Circadian phase was defined by the predicted dim-light melatonin onset (DLMO) and assessed using the BodyTime assay in blood monocytes (*29*). TRE-induced circadian phase changes showed a between-intervention difference with a tendency to the delay for 24 min (95% CI, −5 to 54 min, *P* = 0.10) in lTRE. When comparing samples collected at the end of both interventions, the circadian phase after lTRE was 40 minutes later than that after eTRE (95% CI, 18 to 62 min, *P* = 0.002) (**Fig. 3D**). In agreement with this, the self-reported sleep onset (9 min; 95% CI, 0 to 18 min, *P* = 0.048), offset (20 min; 95% CI, 9 to 32 min, *P* = 0.001), and midpoint (15 min; 95% CI, 7 to 22 min, *P* = 0.001) all occured later in lTRE as compared to eTRE. Sleep duration was not altered by eTRE and lTRE (**Fig. 3E**).

## Discussion

We conducted a randomized, carefully controlled crossover trial comparing 8-hour eTRE vs. lTRE in an intended isocaloric setting. We achieved high adherence to both interventions including successful reductions of eating window under 8 hours (and a prolongation of fasting by about 5 hours) with timely adherence over 96%, unchanged dietary composition and physical activity, which allowed to exclude these potential sources of bias.

The main finding of this study is that neither eTRE nor lTRE improve insulin sensitivity or induce other clinically meaningful changes of cardiometabolic and inflammatory traits in nearly isocaloric conditions. This contradicts our study hypothesis and most published data on TRE, which show beneficial effects on insulin sensitivity (*2, 3, 7, 9, 10, 30*), glucose (*3, 6–8*) and lipid (*6, 8, 11–13*) concentrations, as well as body weight and body fat (*7, 8, 13, 15–17*), whereas eTRE is suggested to be more effective compared to the late or mid-day TRE (*2, 8, 23, 24, 31*). In contrast, long eating windows (*15*), late evening (*15, 32*) and night (*33, 34*) eating, which are common in modern society, are associated with an risk of obesity, diabetes, and other metabolic diseases, at least partly due to the desynchronization of circadian clocks (*35, 36*).

Remarkably, beneficial cardiometabolic effects described previously might be induced by TRE-mediated calorie restriction and not by the shortening of the eating window itself. Indeed, in this nearly isocaloric trial, no improvements in metabolic parameters were observed after two weeks of TRE. Especially, even despite unavoidable small decrease of energy intake in eTRE and minimal weight loss within both interventions (more pronounced in eTRE), insulin sensitivity assessed by Matsuda index, the primary outcome in this trial, showed no between- and within-intervention difference. The post-hoc analysis of another index of insulin sensitivity, OGIS, confirmed this result. This finding aligns with a recent study in subjects with type 2 diabetes, which reported no improvement in insulin sensitivity after three weeks of 10-hour self-selected TRE (*35*). However, it contradicts another study that demonstrated an improvement in insulin sensitivity in 8 men with prediabetes after 5 weeks of eTRE (*2*), although a weight loss was minor in both studies (1.0 kg and 1.4 kg, respectively) and similar to our trial (1.08 kg in eTRE and 0.44 kg in lTRE). Thus, timing or duration of eating window as well as difference in study populations may contribute to the data heterogeneity.

It has to be mentioned that Matsuda insulin sensitivity index, which is calculated using glucose and insulin data from the OGTT, might be compromised by different fasting periods before the OGTT start. In our study, the OGTT was conducted at the same time of day (9:30 hr) to avoid diurnal variation of glucose tolerance (*37, 38*). However, this resulted in a 5-h longer fasting before the OGTT in eTRE compared to lTRE, which can reduce insulin secretion (*39, 40*) and partially explain the decrease in glucose tolerance within eTRE. Indeed, we observed an increase of AUC glucose within eTRE and these changes differed between interventions. This is in agreement with published research (*41*) that estimated an effect size at a 1.7% increase in 2-h plasma glucose for every additional hour of fasting. Further, in our study, we observed a decrease in insulin secretion relative to glucose, as indicated by the insulinogenic index. Thus, the differing fasting durations before the OGTT might mask the true changes (expectedly an increase) in insulin sensitivity assessed by the Matsuda index and potentially explain the absence of observed effects. This also emphasizes the importance of planning for similar fasting durations before OGTT in studies assessing glycemic control, including TRE trials.

Despite the abovementioned OGTT-related bias, clinically relevant outcomes, the mean CGM glucose and other cardiovascular parameters, were not meaningfully affected by eTRE or lTRE. In particular, 24-hour mean glucose concentration, which reflected glycemic control in real-life conditions over 14 days, did not show any differences within or between the TRE interventions. Finally, the absence of TRE-induced changes in most lipid, inflammatory, and oxidative stress markers (as confirmed at the transcriptional level in PBMC) supports the idea that calorie restriction, but not shortening of the eating timing itself, is crucial to induce positive metabolic effects of TRE.

Because our trial observed no effects of TRE on most analyzed parameters in a nearly isocaloric setting, we were unable to compare effects of eTRE and lTRE. When calorie intake is spontaneously reduced, metabolic effects of eTRE are apparently more beneficial compared to lTRE (*3, 7, 10, 17, 19, 21, 42*), but only few trials directly compared eTRE and lTRE (*8, 23, 24*). Xie at al. (*23*) compared early and mid-day TRE in a parallel-arm study and revealed that early eating is more effective for improving of insulin sensitivity, fasting glucose, body mass, and inflammation. Similarly, Zhang et al. (*24*) reported improvements in mean glucose, fasting insulin, and insulin resistance after the eTRE, whereas leptin was reduced after both eTRE and lTRE. Notably, our trial observed similar effects on leptin, confirming TRE’s influence on adipose tissue. In contrast, a single published cross-over study comparing 7-day eTRE and lTRE (*8*) found no difference in improvements in postprandial glucose and fasting triglycerides between the two eating windows. Recent research suggests that the most beneficial eating timing may vary for individuals based on chronotype, genetic, social and other personal factors (*43–45*), highlighting the need for further investigation.

The second key finding of this study suggests that alterations of eating timing shift internal circadian time. We found that shifting the eating window by 5 hours from eTRE to lTRE induces: i) a delay in the circadian phase as estimated by transcript biomarkers in blood monocytes (BodyTime assay), which strongly correlates with DLMO (*29*); ii) expression changes of core clock genes *PER1* and *NR1D1* at a single time point (which can also be caused by the clock phase shift); iii) a delay in sleep timing, which is also controlled by the circadian system (*46*). This finding supports an idea that food intake in humans acts as a *zeitgeber* for circadian clocks as shown in multiple animal studies (*26, 27*) and very few human trials (*28, 47, 48*). The study of Koppold-Liebscher et al. (*28*), which used the same monocyte assay as in our study, observed transient shifts of circadian phase after religious intermittent fasting, characterized by eating at unusual time before sunrise and after the sunset; this effect disappeared 3 months after the return to usual eating time. Wehrens et al. (*47*) demonstrated a delay of PER2 mRNA rhythms in adipose tissue by about one hour after the 5-hour delay in meal times, whereas our study observed a 40-minute shift in samples collected after the lTRE vs. eTRE. Finally, a very recent trial showed that eTRE can advance sleep in late sleepers (*48*). The regulation of peripheral, and possibly also of central, clocks by the timing of food intake may be mediated by postprandial changes of nutrients, metabolites, and hormones (*49*), with insulin apparently playing an essential role (*50*). Taken together, our predicted circadian phase data suggested that eating timing can shift internal circadian clocks in humans, using a single time point sampling. However, to draw a conclusion about whether eating timing-based strategies can resynchronize or restore circadian rhythms in individuals with circadian rhythm disturbances, such as shift workers or people with metabolic diseases, further studies using sampling around the clocks and DLMO assessment should be conducted.

Finally, this trial elucidated an altered hunger regulation by the eating timing during the TRE. In eTRE, subjects felt more hunger, desire and capacity to eat in the morning than in lTRE. This can be explained by longer fasting after the last meal in eTRE at the day before the visit and by a habituation effect to the early eating window, so that the body “expects” the food intake early in the day (*51*). Our study also suggests a hormonal mechanism which might contribute to the hunger regulation by the TRE timing. Whereas the ghrelin concentration did not differ, the morning concentration of anorectic hormone PYY increased within lTRE and declined in eTRE, suggesting a role of PYY in the observed lower hunger in the morning after the lTRE. Whether the eating/fasting timing during TRE alters the circadian rhythm of PYY secretion (*52, 53*) in humans and how it can be related to possible changes in the gut microbiome (*54*) needs to be investigated in the future.

### Limitations

Our study has several limitations which were not mentioned above. Firstly, although the longer intervention period might induce more pronounced metabolic changes, in the ChronoFast study, we decided for the short intervention duration of two weeks. With this, we referred to published TRE trials demonstrated improvement of glycemic control and/or insulin sensitivity even after one or two weeks of intervention (*3, 8, 10*) and our previous isocaloric nutritional trials showed metabolic improvements after similarly short intervention duration (*55, 56*). Moreover, in planning the two-week intervention, we aimed to achieve the maximally accurate 24-hour monitoring of food intake, glucose concentrations, physical activity, and sleep, and to ensure isocaloric conditions through intensive dietary counselling. The short TRE duration aimed to mitigate the spontaneous small reduction in energy intake that, despite all efforts, still occurred within eTRE. This can happen, for example, if subjects skip high-fat or sweet snacks often consumed in the evening. Remarkably, despite this small caloric reduction within eTRE (equivalent to approximately two boiled eggs per day) and the minimal weight loss, we did not observe metabolic improvements in both interventions.

Second limitation is that we enrolled exclusively women to ensure cohort homogeneity, which limits the generalizability of our findings to men. In this context, menstrual cycle changes might potentially confound the results and should be considered. However, this factor is not relevant for our study including only five pre-menopausal and 26 postmenopausal women.

Thirdly, due to COVID-19 hygienic restrictions, we could not use the indirect calorimetry to assess energy expenditure and the BodPod method to measure the body composition. Because the dual energy X-ray absorptiometry (DXA) machine was not available, the assessment of the body composition was conducted by the bioelectrical impedance analysis (BIA) which has some limitations because the BIA accuracy can be affected by factors such as hydration status, food intake, electrode placement, temperature sensitivity, device calibration, and population specificity. However, in our study, the impact of the most abovementioned factors is excluded because of highly standardized measurement conditions, trained staff and homogenous population. In addition, for the percentage of total body water (in relation to body weight), we found no differences within or between interventions (**Table 2**).

Finally, sleep timing assessment was based on the self-reported data using sleep diaries. Nevertheless, although some bias due to subjective assessment is possible, the long-term data collection over 14 days during the baseline and both interventions provides reasonable accuracy of this method.

### Conclusion

In a nearly isocaloric setting, neither eTRE nor lTRE improve insulin sensitivity or other cardiometabolic traits despite changes in the circadian system. This trial highlights the importance of calorie restriction in positive metabolic effects of TRE. Carefully controlled studies should investigate whether the timing of eating influences TRE outcomes in a hypocaloric setting. Further, future research should focus on precision nutrition to identify who will benefit more from eTRE or lTRE interventions.

## Materials and Methods

### Study Design and Randomization

The ChronoFast study was a 10-week randomized crossover trial including two 2-week dietary intervention periods: (1) early time-restricted eating (eTRE) and (2) late time-restricted eating (lTRE), preceded by a 2-4-week baseline (run-in) period and separated by a 2-week washout period (**Fig. 1A**). Participants were randomly allocated to the eTRE-lTRE or lTRE-eTRE study arms based on their BMI and age using the computed minimization method (MinimPy Software) (*57*). Participants visited the clinical study center at the German Institute of Human Nutrition Potsdam-Rehbrücke for the initial screening as well as before and after each dietary intervention (eTRE and lTRE). Visit days began at 8:30 hr and included anthropometrical measurements, fasting blood collection, an OGTT, and nutritional instruction by a dietician. During the 14 days of the baseline and both TRE intervention periods, CGM and actigraphy were performed, and food, sleep and weight diaries were maintained.

The trial was approved by the Ethical Committee of the University of Potsdam, Germany (EA No. 8/2019). All participants provided written informed consent prior to the study participation. The trial protocol was registered at ClinicalTrials.gov (NCT04351672) on April 17, 2020, published previously (*18*). We reported study results using the CONSORT 2025 checklist guideline for reporting randomised trials (*58*).

### Participant Recruitment and Eligibility Criteria

Participants with overweight or obesity from the Berlin-Brandenburg area, Germany, were recruited at the German Institute of Human Nutrition Potsdam-Rehbruecke between April 2020 and December 2021 through flyers, posters, newspaper advertisements, and ads websites. Participants were excluded when they had a male sex; didn’t meet the age range between 18 to 70 years; had a BMI lower than 18 kg/m^2^ or higher than 35 kg/m^2^; did shift work; travelled over more than one time zone in the month prior to the study start; reported weight changes of more than 5% in the past 3 months prior to the study start; were on weight loss medication; were pregnant or breastfeeding; had severe intestinal diseases; practiced special diets, e.g., intermittent fasting; had meaningful sleep disturbances (Pittsburgh Sleep Quality Index (*59*) higher than 10); had diabetes type 1 or 2 or other endocrinological diseases; reported severe renal and liver diseases; reported a stroke the 6 months prior to the study start; had cancer in the 2 years prior to the study start; were on medication with glucocorticoids; had coagulation disorders; were on anticoagulant medication; reported severe anemia or systemic infections; had psychiatric diseases, addictive diseases, or depression. In the case of using other medication or food allergies, the study doctor individually considered whether study participation was possible.

### Time-Restricted Eating Interventions

During the baseline period, the participants followed their usual eating habits, including their habitual eating times. During the TRE interventions, participants had to restrict their eating duration to 8 hours per day but consume their usual kind and amount of food. In the eTRE intervention, participants had to consume food between 8:00 and 16:00 hr, and in the lTRE, between 13:00 and 21:00 hr. During the daily 16-hour fasting period, participants had to consume non-caloric drinks only, such as water, black coffee, or tea, as well as sugar-free chewing gums or mints. Participants were free to divide their meals within the predetermined eating windows. As instruction, and to maintain their habitual daily calorie intake, food composition, and minimize body weight changes, participants received a copy of their individual 14-days food diaries from the baseline period. Within the washout period, participants were requested to return to their habitual eating behavior, including their usual eating window. Participants were also counseled to maintain their habitual physical activity and sleep times throughout the entire trial duration.

### Food Intake and Body Weight Documentation

Food documentation was conducted within the baseline period and both TRE intervention periods for 14 consecutive days. Participants were asked to digitally document their food selection, amount, and time of consumption using the free smartphone app FddB Extender (https://fddb.info/) (*60*). Participants who were not familiar with using a smartphone completed paper-based dietary records, which were then transferred to the FddB Extender app by a study assistance. Prior to the baseline period, all participants received detailed instructions for digital or handwritten food documentation. They were requested to weigh their food whenever possible (or use household measures, e.g., glasses, cups, teaspoons), to add supplemental information (e.g., brand names) and the time of food consumption. The FDDB food database (Fddb Internetportale GmbH, https://fddb.info/) was used to analyse eating timing, energy intake, and macronutrient composition as validated previously (*60*).

Participants were also asked to maintain a constant body weight throughout the study. They were instructed to record their weight daily (in the FddB app or handwritten) and report any weight fluctuation of ≥ 700 g on two consecutive days. Participants were called by the dietician after 7 days of the baseline period and each TRE intervention period to promote adherence to the assigned intervention and answer questions. The FddB Extender app enabled accurate and simple real-time monitoring (*60*) of adherence, including given time frames and potential weight changes. Accessing the nutrition log during the intervention allowed a dietician to intervene rapidly and contact the study subjects in case of adherence problems.

### Assessment of TRE Intervention Adherence

TRE intervention adherence was assessed based on food records using following criteria: 1) duration of the eating window of less than 8 hours; 2) adherence to the prescribed eating time within ± 30 minutes; 3) unchanged energy intake; and 4) unchanged macronutrient composition. For assessment of the duration of eating windows the start of the first calorie intake and end of the last calorie intake were identified in the food records, and the time period between them was assessed for every day of records. The duration of eating window at baseline, eTRE and lTRE was calculated as mean of all days during each study period for each participant. For evaluation of adherence to the prescribed eating times, all days when the subjects followed the required TRE eating time frame with a maximum deviation of ± 30 minutes were considered as compliant. Percentage of compliant days was calculated for each intervention of each participant. Further, energy intake and macronutrient composition (in percentage to daily energy intake) were assessed using the FddB app and FDDB food database for every day of records and mean of all days during the baseline and TRE intervention periods was calculated for each participant.

### Outcome Measures

The primary outcome was insulin sensitivity assessed by the Matsuda index in OGTT. Secondary outcomes were concentrations of glucose and glucose metabolism hormones in OGTT, mean 24-hour glucose, as well as blood pressure, concentrations of lipids, liver enzymes, adipokines, and cytokines, anthropometric parameters, hunger and satiety scores, parameters of intervention adherence, physical activity, and sleep. Exploratory outcomes were the gene expression in PBMC and the internal circadian phase. Outcomes were assessed before and after eTRE and lTRE interventions or during baseline, eTRE, and lTRE periods.

### Anthropometric Measurements, Body Composition, and Blood Pressure

Body weight was measured with a digital scale, and body height was measured with a stadiometer. Waist and hip circumferences were measured using a metric tape. Body composition was assessed by bioimpedance analyzer (BIA; Quantum S, Akern, Florence, Italy) using the method of multi-frequency bioelectrical impedance spectroscopy, with fat and lean mass (in kg) calculated using the related Bodygram^TM^ software (Akern, Italy). Blood pressure was measured on the left upper arm after at least a 3-minute rest, three times, and an average value was calculated. All measurements were performed following an overnight fast.

### Oral Glucose Tolerance Tests and OGTT indices

Fasting blood samples were collected after an overnight fast. For the OGTT, participants consumed a syrup (ACCU-CHEK® Dextrose O.G-T., Roche Diabetes Care, Mannheim, Germany) containing 75g glucose at 9:30 hr within a 5-minute time frame. Blood samples were collected after 30, 60, 90, and 120 minutes using EDTA and serum monovettes (Sarstedt, Nümbrecht, Germany) via an intravenous catheter. To analyse incretins, 10 μg/ml aprotinin (Carl Roth, Karlsruhe, Germany) and 50 μM DPP4 inhibitor (Merck Millipore, Darmstadt, Germany) were added to the blood samples. Serum and plasma samples were centrifuged at 1,800 × g for 10 min at 4°C and stored at −80°C until analysis. In OGTT, indices of insulin sensitivity (Matsuda index), insulin secretion in response to glucose challenge (insulinogenic index), and beta-cell function (disposition index, which characterizes insulin secretion in combination with insulin sensitivity) were calculated based on fasting and postprandial glucose and insulin concentrations using the online calculator at https://mmatsuda.diabetes-smc.jp/MIndex.html. The disposition index is often used as a predictor for the development of type 2 diabetes (*61*). For glucose, free fatty acids, and hormone secretion in OGTT, AUC were determined using the trapezoidal method. In the post hoc analysis, the OGIS index was additionally assessed as described (*62*) using online calculator at http://webmet.pd.cnr.it/ogis/.

### Continuous Glucose Monitoring and Calculation of CGM Indices

The 24h glucose profile was examined using a Freestyle Libre 2 CGM system (Abbott, Wiesbaden, Germany), which measures interstitial fluid glucose with 15 min sampling intervals during the baseline and TRE intervention periods for 14 consequent days. To prevent data loss, participants were instructed to scan the subcutaneous placed sensor on their upper arm with a portable reader. Glycemic control was described by 24-hour mean sensor glucose level (MSG) and the minimum and maximum sensor glucose level. Glycemic variability (GV) was assessed using the EasyGV© software version 9.0.R2 (*63*) (available for non-commercial use at https://www.phc.ox.ac.uk/research/resources/easygv). For this, following intra-day indices were calculated as described previously (*64*): (1) standard deviation (SD) of mean glucose value, indicating variation from the average glucose; (2) mean amplitude of glucose excursions (MAGE), describing the height of glucose excursions above one SD; (3) continuous overlapping net glycemic action (CONGA) as a descriptor for varying glucose concentrations at intervals which were previously set as 60 minutes. Mean of daily differences (MODD), representing the inter-day GV, was calculated as the average of equally timed glucose values across different days. The Easy GV tool also converted glucose data into risk scores, such as the low blood glucose index (LBGI: <0) and high blood glucose index (HBGI: >0). As additional parameter of GV, the coefficient of variation in percentage (CV%) was calculated using the formula SD/MSG x 100 (*64*).

### Biochemical Measurements

Measurement of routine laboratory parameters, i.e., AST, ALT, GGT, hsCRP, glucose, hemoglobin A1c (HbA1c), total cholesterol, HDL cholesterol, and non-esterified free fatty acids, were performed using ABX Pentra (HORIBA ABX SAS, Montpellier, France). LDL cholesterol was determined using the Friedewald equation. Circulating concentrations of insulin (10-1113-01), C-peptide (10-1136-01) and glucagon (10-1271-01, all from Mercodia Inc., NC, USA), MCP-1 (DCP00, Quantikine®ELISA, R&D Systems Inc., MN, USA), IL-6 (HS600C) and TNF-α (HSTA00E, both Quantikine®ELISA HS, R&D Systems Inc., MN, USA), PYY (EZHPYYT66K, Merck Millipore, Darmstadt, Germany), adiponectin (RD195023100, Biovendor, Germany), and leptin (DLP00, Quantikine®ELISA, R&D Systems Inc., MN, USA) were quantified using commercial enzyme-linked immunosorbent assay (ELISA) kits using a DSX® 4-Plate ELISA Processing System (DYNEX Technologies GmbH, Denkendorf, Germany). Ghrelin ELISA (RA194063500R, Biovendor, Heidelberg, Germany) was conducted using a BioTek Eon™ plate reader (BioTek Instruments, Bad Friedrichshall, Germany).

### Oxidative Stress Markers

Malondialdehyde (MDA) was measured in plasma samples after derivatization with thiobarbituric acid (TBA) and separation by reverse-phase high performance liquid chromatography (HPLC) coupled with fluorescence detection. 3-Nitrotyrosine and protein carbonyls were assessed by indirect in-house ELISA with nitrated bovine serum albumin (BSA) and oxidized BSA as standards, respectively, as previously described (*65*).

### Assessment of Physical Activity and Metabolic Equivalent

Participants were asked to maintain their habitual physical activity throughout the entire trial duration. 24-hour physical activity was monitored using a blinded accelerometer (ActiGraph wGT3X-BT, ActiGraph Corporation, Pensacola, FL, USA), which was placed on the wrist of the non-dominant arm for 14 days during the baseline phase and both TRE interventions. The device was to be removed only during bathing or swimming, and participants were asked to record non-wear times in a protocol. The analysis of energy expenditure, physical activity levels, and the metabolic equivalent of task (MET) was conducted using ActiLife software version 6.13.4 (ActiGraph Corporation, Pensacola, FL, USA).

### Chronotype assessment and Sleep Timing

Individual chronotypes were assessed using the Munich Chronotype Questionnaire (MCTQ) (*66*) and the Horne-Östberg Morningness-Eveningness Questionnaire (MEQ) (*67*). For chronotype classification, the MSFsc value (midsleep on free days corrected for the sleep debt over the working days) was used. MCF-Sc <4 was defined as an early chronotype, MCF-Sc >5 as a late chronotype, and intermediate values as a normal chronotype. Classification in MEQ also based on standardized criteria with a range from 59-69 for early chronotypes, 31-41 for late chronotypes, and intermediate values for normal chronotypes. Extreme values in MEQ outside the mentioned ranges were counted as early and late chronotypes and not classified as extreme chronotypes. In case of heterogenous results in both questionnaires, the MCTQ was used for the final chronotype classification. Participants were asked to maintain their habitual sleep times throughout the trial. Sleep timing (sleep onset and offset) was monitored using a sleep diary for 14 days during the baseline and both intervention periods.

### Hunger and Satiety Scores

The assessment of hunger and satiety was conducted on the last day of both TRE-intervention periods in the morning (at 8:00 hr) and in the evening (at 20:00 hr) using a Visual Analog Scale (VAS). The questionnaire included four questions about (i) the desire to eat, (ii) hunger, (iii) satiety, and (iv) the capacity to eat.

### Gene Expression and Circadian Phase in Blood Monocytes

Peripheral blood mononuclear cells (PBMC) were isolated from fasting EDTA blood collected between 8:30 and 11:00 hr using BD Vacutainer® CPT with an integrated FICOLL™ gradient (BD Biosciences, Heidelberg, Germany). For gene expression analysis, total RNA was purified using the NucleoSpin RNA midi kit (Macherey-Nagel, Berlin, Germany). RNA concentration was measured using a NanoDrop ND-1000 spectrophotometer (Peqlab Biotechnologie, Erlangen, Germany). Single-stranded cDNA was synthesized using the high-capacity cDNA reverse transcription kit (Applied Biosystems™, Carlsbad, USA). Quantitative real-time PCR (qPCR) was performed with the ViiA 7 sequence detection system using Power SYBR Green PCR Master Mix (Applied Biosystems, Foster City, CA, USA) and specific primers (**Table S2**). Gene expression was assessed by the standard curve method and normalized to the housekeeper gene beta-2-microglobulin (B2M).

The circadian phase was defined by DLMO, which typically occurs about two hours before habitual bedtime, and was assessed using the recently validated BodyTime assay. This assay requires only a single blood sample to objectively and accurately determine the phase of the circadian clock of an individual as validated previously (*29*). For the analysis, blood monocytes were purified from PBMC samples using CD14 microbeads (Miltenyi Biotech, Bergisch Gladbach, Germany) via positive magnetic sorting. Total RNA was isolated from monocytes using TRIzol reagent (Invitrogen, Karlsruhe, Germany). 1 µg RNA was used to analyze the expression of 20 biomarker genes using the NanoString nCounter platform (NanoString Technologies, Seattle, WA, USA) as described (*29*).

### Sample Size Calculation

Power calculation was completed using the G-Power software v.3.1 (*68*) for the primary end point of change in insulin sensitivity. The calculation was based on the difference in insulin sensitivity observed in the meal tolerance tests conducted in the morning and in the evening in subjects with IFG/IGT in our previous study (*37*). The trial was statistically powered to detect a difference in insulin sensitivity of 12.8 % plus or minus 7.1 % at a significance level of 0.05 and a statistical power of 80%. Assuming a 10% dropout rate, an estimated 33 enrollees were needed to ensure that 30 individuals completed the trial. Recruitment stopped once the target sample size was exceeded (n=31).

### Blinding

Participants and intervention staff were unblinded. Prior to randomization, all outcomes were collected blinded. About 70% of the post-intervention data were assessed blinded. Specifically, all blood parameters, CGM, physical activity, and gene expression measurements were performed blinded, whereas anthropometric measurements, food intake, and chronotype assessments were not. The dietician who analyzed food records and estimated TRE adherence was not blinded to group assignments. Other investigators and statisticians were blinded during the study procedures and were unblinded only after all data had been analyzed.

### Statistical Methods

Data analyses were performed using SPSS 28.0 software (SPSS, Chicago, IL, USA) using 2-sided tests at α = 0.05. All analyses were intention-to-treat. All data were initially checked for missing values, cleaned, and inspected to determine ranges, identify outliers, and assess data distribution using the Shapiro–Wilk test. Absolute values were expressed as mean ± SD when normally distributed and median (IQR) when not normally distributed. Changes within-intervention and differences between-intervention were expressed as mean (95% CI). Between-intervention (parameter changes in lTRE vs. changes in eTRE) comparisons for anthropometric parameters and biochemical measurements were assessed by linear mixed models. The model included cardiometabolic parameters as dependent variables, treatment (eTRE or lTRE), period (first or second) and residual effect of the first experimental period over the second period (carryover effect) as fixed factors, and subjects as a random factor (*37*). In the whole cohort, four outcomes - waist circumference, AUC glucagon, AST, and 3-nitrotyrosine - had significant period effects, which are reported in table and figure legends; all other outcomes did not. Total cholesterol showed a carryover effect and was therefore analyzed for the first intervention only. Within-intervention comparisons (values after/during the intervention vs. values before the intervention) and between-intervention (parameter changes in lTRE vs. changes in eTRE) comparisons for all other outcomes were assessed using paired Student’s t-test for normally distributed data or the Wilcoxon test for non-normally distributed data. Comparisons of two independent groups (values in NGT vs. IFG/IGT groups) were conducted using Student’s unpaired t-test for normally distributed data or the Mann–Whitney U-test for non-normally distributed data. Our analyses did not adjust p-values for multiple comparisons when analyzing cardiometabolic outcomes, which is consistent with a majority of published nutritional and weight loss clinical trials. The visualization of the data was performed using GraphPad Prism software version 10.2.3 (GraphPad Prism Inc., La Jolla, CA, USA).

## LIST OF SUPPLEMENTARY MATERIALS

The PDF file includes:

Fig. S1 to S4 Tables S1 to S8

## Data Availability

All data produced in the present study are available upon reasonable request to the authors

## Acknowledgements

We acknowledge all study participants for their cooperation. We gratefully thank Manuela Bergmann for the support in the clinical trial preparation, Juliane Roeder, Melanie Hannemann, and Lothar Napieralski for their work with study subjects, Katja Treu and Christiana Gerbracht for the help in the preparation of nutritional counselling, Marion Urbich and Nadine Huckauf for excellent technical assistance in the sample analysis. We also thank all students contributed to the clinical trial and Maya Patterson for language proof-reading of the manuscript.

## Author contributions

Conceptualization: A.M., D.A.K., A.F.H.P., A.K., and O.P.-R; Methodology: A.M., D.A.K., A.F.H.P., A.K., and O.P.-R.; Software: N.S., B.A.; Validation: B.P., B.S., J.S., and A.O.; Formal analysis: B.P., B.S., J.S., N.S., A.K., and O.P.-R.; Investigation: B.P., B.S., J.S., D.W., and A.K.; Resources: A.M., D.A.K, N.S., B.A., K.M., T.G., A.K., and O.P.-R.; Data curation: B.P., B.S., J.S., N.S.; Writing - original draft: B.P. and O.P.-R; Writing - review & editing: All authors; Visualization: B.P. and O.P.-R; Supervision: O.P.-R.; Project administration: O.P.-R.; Funding acquisition: O.P.-R.

## Data and materials availability

All data associated with this study are present in the paper or the Supplementary Materials. This trial is registered in ClinicalTrials.gov, NCT04351672. This work is licensed under a Creative Commons Attribution 4.0 International (CC BY 4.0) license, which permits unrestricted use, distribution, and reproduction in any medium, provided the original work is properly cited. To view a copy of this license, visit http://creativecommons.org/licenses/by/4.0/. This license does not apply to figures/photos/artwork or other content included in the article that is credited to a third party; obtain authorization from the rights holder before using this material.

## References

1. D. A. Koppold, C. Breinlinger, E. Hanslian, C. Kessler, H. Cramer, A. R. Khokhar, C. M. Peterson, G. Tinsley, C. Vernieri, R. J. Bloomer, M. Boschmann, N. L. Bragazzi, S. Brandhorst, K. Gabel, A. C. Goldhamer, M. M. Grajower, M. Harvie, L. Heilbronn, B. D. Horne, S. N. Karras, J. Langhorst, E. Lischka, F. Madeo, S. J. Mitchell, I. E. Papagiannopoulos-Vatopaidinos, M. Papagiannopoulou, H. Pijl, E. Ravussin, M. Ritzmann-Widderich, K. Varady, L. Adamidou, M. Chihaoui, R. de Cabo, M. Hassanein, N. Lessan, V. Longo, E. N. C. Manoogian, M. P. Mattson, J. B. Muhlestein, S. Panda, S. K. Papadopoulou, N. E. Rodopaios, R. Stange, A. Michalsen, International consensus on fasting terminology. Cell metabolism 36, 1779–1794 e1774 (2024); published online EpubAug 6 (10.1016/j.cmet.2024.06.013).

2. E. F. Sutton, R. Beyl, K. S. Early, W. T. Cefalu, E. Ravussin, C. M. Peterson, Early Time-Restricted Feeding Improves Insulin Sensitivity, Blood Pressure, and Oxidative Stress Even without Weight Loss in Men with Prediabetes. Cell Metab 27, 1212–1221.e1213 (2018); published online EpubJun 5 (10.1016/j.cmet.2018.04.010).

3. H. Jamshed, R. A. Beyl, D. L. Della Manna, E. S. Yang, E. Ravussin, C. M. Peterson, Early Time-Restricted Feeding Improves 24-Hour Glucose Levels and Affects Markers of the Circadian Clock, Aging, and Autophagy in Humans. Nutrients 11, (2019); published online EpubMay 30 (10.3390/nu11061234).

4. A. Chaix, A. Zarrinpar, P. Miu, S. Panda, Time-restricted feeding is a preventative and therapeutic intervention against diverse nutritional challenges. Cell Metab 20, 991–1005 (2014); published online EpubDec 2 (10.1016/j.cmet.2014.11.001).

5. M. Hatori, C. Vollmers, A. Zarrinpar, L. DiTacchio, E. A. Bushong, S. Gill, M. Leblanc, A. Chaix, M. Joens, J. A. Fitzpatrick, M. H. Ellisman, S. Panda, Time-restricted feeding without reducing caloric intake prevents metabolic diseases in mice fed a high-fat diet. Cell metabolism 15, 848–860 (2012); published online EpubJun 6 (10.1016/j.cmet.2012.04.019).

6. L. S. Chow, E. N. C. Manoogian, A. Alvear, J. G. Fleischer, H. Thor, K. Dietsche, Q. Wang, J. S. Hodges, N. Esch, S. Malaeb, T. Harindhanavudhi, K. S. Nair, S. Panda, D. G. Mashek, Time-Restricted Eating Effects on Body Composition and Metabolic Measures in Humans who are Overweight: A Feasibility Study. Obesity (Silver Spring) 28, 860–869 (2020); published online EpubMay (10.1002/oby.22756).

7. T. Moro, G. Tinsley, A. Bianco, G. Marcolin, Q. F. Pacelli, G. Battaglia, A. Palma, P. Gentil, M. Neri, A. Paoli, Effects of eight weeks of time-restricted feeding (16/8) on basal metabolism, maximal strength, body composition, inflammation, and cardiovascular risk factors in resistance-trained males. J Transl Med 14, 290 (2016); published online EpubOct 13 (10.1186/s12967-016-1044-0).

8. A. T. Hutchison, P. Regmi, E. N. C. Manoogian, J. G. Fleischer, G. A. Wittert, S. Panda, L. K. Heilbronn, Time-Restricted Feeding Improves Glucose Tolerance in Men at Risk for Type 2 Diabetes: A Randomized Crossover Trial. Obesity (Silver Spring) 27, 724–732 (2019); published online EpubMay (10.1002/oby.22449).

9. S. Cienfuegos, K. Gabel, F. Kalam, M. Ezpeleta, E. Wiseman, V. Pavlou, S. Lin, M. L. Oliveira, K. A. Varady, Effects of 4- and 6-h Time-Restricted Feeding on Weight and Cardiometabolic Health: A Randomized Controlled Trial in Adults with Obesity. Cell Metab 32, 366–378.e363 (2020); published online EpubSep 1 (10.1016/j.cmet.2020.06.018).

10. R. Jones, P. Pabla, J. Mallinson, A. Nixon, T. Taylor, A. Bennett, K. Tsintzas, Two weeks of early time-restricted feeding (eTRF) improves skeletal muscle insulin and anabolic sensitivity in healthy men. Am J Clin Nutr 112, 1015–1028 (2020); published online EpubOct 1 (10.1093/ajcn/nqaa192).

11. H. Cai, Y. L. Qin, Z. Y. Shi, J. H. Chen, M. J. Zeng, W. Zhou, R. Q. Chen, Z. Y. Chen, Effects of alternate-day fasting on body weight and dyslipidaemia in patients with non-alcoholic fatty liver disease: a randomised controlled trial. BMC Gastroenterol 19, 219 (2019); published online EpubDec 18 (10.1186/s12876-019-1132-8).

12. F. Zeb, X. Wu, L. Chen, S. Fatima, I. U. Haq, A. Chen, F. Majeed, Q. Feng, M. Li, Effect of time-restricted feeding on metabolic risk and circadian rhythm associated with gut microbiome in healthy males. Br J Nutr 123, 1216–1226 (2020); published online EpubJun 14 (10.1017/s0007114519003428).

13. M. J. Wilkinson, E. N. C. Manoogian, A. Zadourian, H. Lo, S. Fakhouri, A. Shoghi, X. Wang, J. G. Fleischer, S. Navlakha, S. Panda, P. R. Taub, Ten-Hour Time-Restricted Eating Reduces Weight, Blood Pressure, and Atherogenic Lipids in Patients with Metabolic Syndrome. Cell Metab 31, 92–104 e105 (2020); published online EpubJan 7 (10.1016/j.cmet.2019.11.004).

14. K. Gabel, J. Marcell, K. Cares, F. Kalam, S. Cienfuegos, M. Ezpeleta, K. A. Varady, Effect of time restricted feeding on the gut microbiome in adults with obesity: A pilot study. Nutr Health 26, 79–85 (2020); published online EpubJun (10.1177/0260106020910907).

15. S. Gill, S. Panda, A Smartphone App Reveals Erratic Diurnal Eating Patterns in Humans that Can Be Modulated for Health Benefits. Cell metabolism 22, 789–798 (2015); published online EpubNov 3 (10.1016/j.cmet.2015.09.005).

16. K. Gabel, K. K. Hoddy, N. Haggerty, J. Song, C. M. Kroeger, J. F. Trepanowski, S. Panda, K. A. Varady, Effects of 8-hour time restricted feeding on body weight and metabolic disease risk factors in obese adults: A pilot study. Nutr Healthy Aging 4, 345–353 (2018); published online EpubJun 15 (10.3233/NHA-170036).

17. S. Cienfuegos, K. Gabel, F. Kalam, M. Ezpeleta, E. Wiseman, V. Pavlou, S. Lin, M. L. Oliveira, K. A. Varady, Effects of 4- and 6-h Time-Restricted Feeding on Weight and Cardiometabolic Health: A Randomized Controlled Trial in Adults with Obesity. Cell metabolism 32, 366–378 e363 (2020); published online EpubSep 1 (10.1016/j.cmet.2020.06.018).

18. B. Peters, D. A. Koppold-Liebscher, B. Schuppelius, N. Steckhan, A. F. H. Pfeiffer, A. Kramer, A. Michalsen, O. Pivovarova-Ramich, Effects of Early vs. Late Time-Restricted Eating on Cardiometabolic Health, Inflammation, and Sleep in Overweight and Obese Women: A Study Protocol for the ChronoFast Trial. Front Nutr 8, 765543 (2021)10.3389/fnut.2021.765543).

19. B. Schuppelius, B. Peters, A. Ottawa, O. Pivovarova-Ramich, Time Restricted Eating: A Dietary Strategy to Prevent and Treat Metabolic Disturbances. Frontiers in endocrinology 12, 683140 (2021)10.3389/fendo.2021.683140).

20. S. Bhasin, Time-Restricted Eating to Improve Health-A Promising Idea in Need of Stronger Clinical Trial Evidence. JAMA Intern Med 182, 963–964 (2022); published online EpubSep 1 (10.1001/jamainternmed.2022.3038).

21. E. Ravussin, R. A. Beyl, E. Poggiogalle, D. S. Hsia, C. M. Peterson, Early Time-Restricted Feeding Reduces Appetite and Increases Fat Oxidation But Does Not Affect Energy Expenditure in Humans. Obesity (Silver Spring) 27, 1244–1254 (2019); published online EpubAug (10.1002/oby.22518).

22. E. Poggiogalle, H. Jamshed, C. M. Peterson, Circadian regulation of glucose, lipid, and energy metabolism in humans. Metabolism: clinical and experimental 84, 11–27 (2018); published online EpubJul (10.1016/j.metabol.2017.11.017).

23. Z. Xie, Y. Sun, Y. Ye, D. Hu, H. Zhang, Z. He, H. Zhao, H. Yang, Y. Mao, Randomized controlled trial for time-restricted eating in healthy volunteers without obesity. Nat Commun 13, 1003 (2022); published online EpubFeb 22 (10.1038/s41467-022-28662-5).

24. L. M. Zhang, Z. Liu, J. Q. Wang, R. Q. Li, J. Y. Ren, X. Gao, S. S. Lv, L. Y. Liang, F. Zhang, B. W. Yin, Y. Sun, H. Tian, H. C. Zhu, Y. T. Zhou, Y. X. Ma, Randomized controlled trial for time-restricted eating in overweight and obese young adults. iScience 25, 104870 (2022); published online EpubSep 16 (10.1016/j.isci.2022.104870).

25. S. Panda, Circadian physiology of metabolism. Science 354, 1008–1015 (2016); published online EpubNov 25 (10.1126/science.aah4967).

26. F. Damiola, N. Le Minh, N. Preitner, B. Kornmann, F. Fleury-Olela, U. Schibler, Restricted feeding uncouples circadian oscillators in peripheral tissues from the central pacemaker in the suprachiasmatic nucleus. Genes Dev 14, 2950–2961 (2000); published online EpubDec 1 (10.1101/gad.183500).

27. A. Mukherji, A. Kobiita, M. Damara, N. Misra, H. Meziane, M. F. Champy, P. Chambon, Shifting eating to the circadian rest phase misaligns the peripheral clocks with the master SCN clock and leads to a metabolic syndrome. Proceedings of the National Academy of Sciences of the United States of America 112, E6691–6698 (2015); published online EpubDec 01 (10.1073/pnas.1519807112).

28. D. A. Koppold-Liebscher, C. Klatte, S. Demmrich, J. Schwarz, F. I. Kandil, N. Steckhan, R. Ring, C. S. Kessler, M. Jeitler, B. Koller, B. Ananthasubramaniam, C. Eisenmann, A. Mähler, M. Boschmann, A. Kramer, A. Michalsen, Effects of Daytime Dry Fasting on Hydration, Glucose Metabolism and Circadian Phase: A Prospective Exploratory Cohort Study in Bahá’í Volunteers. Front Nutr 8, 662310 (2021)10.3389/fnut.2021.662310).

29. N. Wittenbrink, B. Ananthasubramaniam, M. Munch, B. Koller, B. Maier, C. Weschke, F. Bes, J. de Zeeuw, C. Nowozin, A. Wahnschaffe, S. Wisniewski, M. Zaleska, O. Bartok, R. Ashwal-Fluss, H. Lammert, H. Herzel, M. Hummel, S. Kadener, D. Kunz, A. Kramer, High-accuracy determination of internal circadian time from a single blood sample. The Journal of clinical investigation 128, 3826–3839 (2018); published online EpubAug 31 (10.1172/JCI120874).

30. S. Cienfuegos, K. Gabel, F. Kalam, M. Ezpeleta, V. Pavlou, S. Lin, E. Wiseman, K. A. Varady, The effect of 4-h versus 6-h time restricted feeding on sleep quality, duration, insomnia severity and obstructive sleep apnea in adults with obesity. Nutr Health 28, 5–11 (2022); published online EpubMar (10.1177/02601060211002347).

31. H. Jamshed, F. L. Steger, D. R. Bryan, J. S. Richman, A. H. Warriner, C. J. Hanick, C. K. Martin, S. J. Salvy, C. M. Peterson, Effectiveness of Early Time-Restricted Eating for Weight Loss, Fat Loss, and Cardiometabolic Health in Adults With Obesity: A Randomized Clinical Trial. JAMA Intern Med 182, 953–962 (2022); published online EpubSep 1 (10.1001/jamainternmed.2022.3050).

32. N. J. Gupta, V. Kumar, S. Panda, A camera-phone based study reveals erratic eating pattern and disrupted daily eating-fasting cycle among adults in India. PloS one 12, e0172852 (2017)10.1371/journal.pone.0172852).

33. L. C. Antunes, R. Levandovski, G. Dantas, W. Caumo, M. P. Hidalgo, Obesity and shift work: chronobiological aspects. Nutr Res Rev 23, 155–168 (2010); published online EpubJun (10.1017/S0954422410000016).

34. F. A. Scheer, M. F. Hilton, C. S. Mantzoros, S. A. Shea, Adverse metabolic and cardiovascular consequences of circadian misalignment. Proc Natl Acad Sci U S A 106, 4453–4458 (2009); published online EpubMar 17 (10.1073/pnas.0808180106).

35. C. Andriessen, C. E. Fealy, A. Veelen, S. M. M. van Beek, K. H. M. Roumans, N. J. Connell, J. Mevenkamp, E. Moonen-Kornips, B. Havekes, V. B. Schrauwen-Hinderling, J. Hoeks, P. Schrauwen, Three weeks of time-restricted eating improves glucose homeostasis in adults with type 2 diabetes but does not improve insulin sensitivity: a randomised crossover trial. Diabetologia 65, 1710–1720 (2022); published online EpubOct (10.1007/s00125-022-05752-z).

36. M. Franzago, E. Alessandrelli, S. Notarangelo, L. Stuppia, E. Vitacolonna, Chrono-Nutrition: Circadian Rhythm and Personalized Nutrition. Int J Mol Sci 24, (2023); published online EpubJan 29 (10.3390/ijms24032571).

37. K. Kessler, S. Hornemann, K. J. Petzke, M. Kemper, A. Kramer, A. F. Pfeiffer, O. Pivovarova, N. Rudovich, The effect of diurnal distribution of carbohydrates and fat on glycaemic control in humans: a randomized controlled trial. Sci Rep 7, 44170 (2017); published online EpubMar 8 (10.1038/srep44170).

38. A. Saad, C. Dalla Man, D. K. Nandy, J. A. Levine, A. E. Bharucha, R. A. Rizza, R. Basu, R. E. Carter, C. Cobelli, Y. C. Kudva, A. Basu, Diurnal pattern to insulin secretion and insulin action in healthy individuals. Diabetes 61, 2691–2700 (2012); published online EpubNov (10.2337/db11-1478).

39. S. W. Jorgensen, L. Hjort, L. Gillberg, L. Justesen, S. Madsbad, C. Brons, A. A. Vaag, Impact of prolonged fasting on insulin secretion, insulin action, and hepatic versus whole body insulin secretion disposition indices in healthy young males. American journal of physiology. Endocrinology and metabolism 320, E281–E290 (2021); published online EpubFeb 1 (10.1152/ajpendo.00433.2020).

40. A. Nas, N. Mirza, F. Hägele, J. Kahlhöfer, J. Keller, R. Rising, T. A. Kufer, A. Bosy-Westphal, Impact of breakfast skipping compared with dinner skipping on regulation of energy balance and metabolic risk. Am J Clin Nutr 105, 1351–1361 (2017); published online EpubJun (10.3945/ajcn.116.151332).

41. K. K. B. Clemmensen, J. S. Quist, D. Vistisen, D. R. Witte, A. Jonsson, O. Pedersen, T. Hansen, J. J. Holst, T. Lauritzen, M. E. Jørgensen, S. Torekov, K. Færch, Role of fasting duration and weekday in incretin and glucose regulation. Endocr Connect 9, 279–288 (2020); published online EpubApr (10.1530/ec-20-0009).

42. D. A. Lowe, N. Wu, L. Rohdin-Bibby, A. H. Moore, N. Kelly, Y. E. Liu, E. Philip, E. Vittinghoff, S. B. Heymsfield, J. E. Olgin, J. A. Shepherd, E. J. Weiss, Effects of Time-Restricted Eating on Weight Loss and Other Metabolic Parameters in Women and Men With Overweight and Obesity: The TREAT Randomized Clinical Trial. JAMA Intern Med 180, 1491–1499 (2020); published online EpubNov 1 (10.1001/jamainternmed.2020.4153).

43. A. W. McHill, A. J. Phillips, C. A. Czeisler, L. Keating, K. Yee, L. K. Barger, M. Garaulet, F. A. Scheer, E. B. Klerman, Later circadian timing of food intake is associated with increased body fat. Am J Clin Nutr 106, 1213–1219 (2017); published online EpubNov (10.3945/ajcn.117.161588).

44. M. Garaulet, J. Lopez-Minguez, H. S. Dashti, C. Vetter, A. M. Hernandez-Martinez, M. Perez-Ayala, J. C. Baraza, W. Wang, J. C. Florez, F. Scheer, R. Saxena, Interplay of Dinner Timing and MTNR1B Type 2 Diabetes Risk Variant on Glucose Tolerance and Insulin Secretion: A Randomized Crossover Trial. Diabetes care 45, 512–519 (2022); published online EpubMar 1 (10.2337/dc21-1314).

45. B. Peters, J. Vahlhaus, O. Pivovarova-Ramich, Meal timing and its role in obesity and associated diseases. Frontiers in endocrinology 15, 1359772 (2024)10.3389/fendo.2024.1359772).

46. L. H. Ashbrook, A. D. Krystal, Y. H. Fu, L. J. Ptáček, Genetics of the human circadian clock and sleep homeostat. Neuropsychopharmacology 45, 45–54 (2020); published online EpubJan (10.1038/s41386-019-0476-7).

47. S. M. T. Wehrens, S. Christou, C. Isherwood, B. Middleton, M. A. Gibbs, S. N. Archer, D. J. Skene, J. D. Johnston, Meal Timing Regulates the Human Circadian System. Current biology: CB 27, 1768–1775 e1763 (2017); published online EpubJun 19 (10.1016/j.cub.2017.04.059).

48. D. J. Blum, B. Hernandez, J. M. Zeitzer, Early time-restricted eating advances sleep in late sleepers: a pilot randomized controlled trial. J Clin Sleep Med 19, 2097–2106 (2023); published online EpubDec 1 (10.5664/jcsm.10754).

49. A. Chaudhari, R. Gupta, K. Makwana, R. Kondratov, Circadian clocks, diets and aging. Nutr Healthy Aging 4, 101–112 (2017); published online EpubMar 31 (10.3233/NHA-160006).

50. N. Tuvia, O. Pivovarova-Ramich, V. Murahovschi, S. Luck, A. Grudziecki, A. C. Ost, M. Kruse, V. J. Nikiforova, M. Osterhoff, P. Gottmann, O. Gogebakan, C. Sticht, N. Gretz, M. Schupp, A. Schurmann, N. Rudovich, A. F. H. Pfeiffer, A. Kramer, Insulin directly regulates the circadian clock in adipose tissue. Diabetes, (2021); published online EpubJul 5 (10.2337/db20-0910).

51. L. H. Epstein, K. A. Carr, Food reinforcement and habituation to food are processes related to initiation and cessation of eating. Physiology & behavior 239, 113512 (2021); published online EpubOct 1 (10.1016/j.physbeh.2021.113512).

52. C. A. Rynders, S. J. Morton, D. H. Bessesen, K. P. Wright, Jr., J. L. Broussard, Circadian Rhythm of Substrate Oxidation and Hormonal Regulators of Energy Balance. Obesity (Silver Spring) 28 Suppl 1, S104–S113 (2020); published online EpubJul (10.1002/oby.22816).

53. M. J. Maroni, K. M. Capri, A. V. Cushman, H. V. Deane, H. Concepcion, H. DeCourcey, J. A. Seggio, The timing of fasting leads to different levels of food consumption and PYY(3-36) in nocturnal mice. Hormones (Athens) 19, 549–558 (2020); published online EpubDec (10.1007/s42000-020-00221-x).

54. E. S. Chambers, A. Viardot, A. Psichas, D. J. Morrison, K. G. Murphy, S. E. Zac-Varghese, K. MacDougall, T. Preston, C. Tedford, G. S. Finlayson, J. E. Blundell, J. D. Bell, E. L. Thomas, S. Mt-Isa, D. Ashby, G. R. Gibson, S. Kolida, W. S. Dhillo, S. R. Bloom, W. Morley, S. Clegg, G. Frost, Effects of targeted delivery of propionate to the human colon on appetite regulation, body weight maintenance and adiposity in overweight adults. Gut 64, 1744–1754 (2015); published online EpubNov (10.1136/gutjnl-2014-307913).

55. S. Sucher, M. Markova, S. Hornemann, O. Pivovarova, N. Rudovich, R. Thomann, R. Schneeweiss, S. Rohn, A. F. H. Pfeiffer, Comparison of the effects of diets high in animal or plant protein on metabolic and cardiovascular markers in type 2 diabetes: A randomized clinical trial. Diabetes Obes Metab 19, 944–952 (2017); published online EpubJul (10.1111/dom.12901).

56. T. Frahnow, M. A. Osterhoff, S. Hornemann, M. Kruse, M. A. Surma, C. Klose, K. Simons, A. F. H. Pfeiffer, Heritability and responses to high fat diet of plasma lipidomics in a twin study. Sci Rep 7, 3750 (2017); published online EpubJun 16 (10.1038/s41598-017-03965-6).

57. M. Saghaei, An overview of randomization and minimization programs for randomized clinical trials. J Med Signals Sens 1, 55–61 (2011); published online EpubJan (

58. S. Hopewell, A.-W. Chan, G. S. Collins, A. Hróbjartsson, D. Moher, K. F. Schulz, R. Tunn, R. Aggarwal, M. Berkwits, J. A. Berlin, N. Bhandari, N. J. Butcher, M. K. Campbell, R. C. W. Chidebe, D. Elbourne, A. Farmer, D. A. Fergusson, R. M. Golub, S. N. Goodman, T. C. Hoffmann, J. P. A. Ioannidis, B. C. Kahan, R. L. Knowles, S. E. Lamb, S. Lewis, E. Loder, M. Offringa, P. Ravaud, D. P. Richards, F. W. Rockhold, D. L. Schriger, N. L. Siegfried, S. Staniszewska, R. S. Taylor, L. Thabane, D. Torgerson, S. Vohra, I. R. White, I. Boutron, CONSORT 2025 statement: updated guideline for reporting randomised trials. BMJ 389, e081123 (2025)10.1136/bmj-2024-081123).

59. D. J. Buysse, C. F. Reynolds, 3rd, T. H. Monk, S. R. Berman, D. J. Kupfer, The Pittsburgh Sleep Quality Index: a new instrument for psychiatric practice and research. Psychiatry Res 28, 193–213 (1989); published online EpubMay (10.1016/0165-1781(89)90047-4).

60. I. Baum Martinez, B. Peters, J. Schwarz, B. Schuppelius, N. Steckhan, D. A. Koppold-Liebscher, A. Michalsen, O. Pivovarova-Ramich, Validation of a Smartphone Application for the Assessment of Dietary Compliance in an Intermittent Fasting Trial. Nutrients 14, (2022); published online EpubSep 7 (10.3390/nu14183697).

61. K. M. Utzschneider, R. L. Prigeon, M. V. Faulenbach, J. Tong, D. B. Carr, E. J. Boyko, D. L. Leonetti, M. J. McNeely, W. Y. Fujimoto, S. E. Kahn, Oral disposition index predicts the development of future diabetes above and beyond fasting and 2-h glucose levels. Diabetes care 32, 335–341 (2009); published online EpubFeb (10.2337/dc08-1478).

62. A. Mari, G. Pacini, E. Murphy, B. Ludvik, J. J. Nolan, A model-based method for assessing insulin sensitivity from the oral glucose tolerance test. Diabetes Care 24, 539–548 (2001); published online EpubMar (10.2337/diacare.24.3.539).

63. N. R. Hill, N. S. Oliver, P. Choudhary, J. C. Levy, P. Hindmarsh, D. R. Matthews, Normal reference range for mean tissue glucose and glycemic variability derived from continuous glucose monitoring for subjects without diabetes in different ethnic groups. Diabetes technology & therapeutics 13, 921–928 (2011); published online EpubSep (10.1089/dia.2010.0247).

64. C. L. Pappe, B. Peters, H. Dommisch, J. P. Woelber, O. Pivovarova-Ramich, Effects of reducing free sugars on 24-hour glucose profiles and glycemic variability in subjects without diabetes. Front Nutr 10, 1213661 (2023)10.3389/fnut.2023.1213661).

65. D. Weber, W. Stuetz, W. Bernhard, A. Franz, M. Raith, T. Grune, N. Breusing, Oxidative stress markers and micronutrients in maternal and cord blood in relation to neonatal outcome. Eur J Clin Nutr 68, 215–222 (2014); published online EpubFeb (10.1038/ejcn.2013.263).

66. T. Roenneberg, A. Wirz-Justice, M. Merrow, Life between clocks: daily temporal patterns of human chronotypes. J Biol Rhythms 18, 80–90 (2003); published online EpubFeb (10.1177/0748730402239679).

67. J. A. Horne, O. Ostberg, A self-assessment questionnaire to determine morningness-eveningness in human circadian rhythms. Int J Chronobiol 4, 97–110 (1976).

68. F. Faul, E. Erdfelder, A. G. Lang, A. Buchner, G*Power 3: a flexible statistical power analysis program for the social, behavioral, and biomedical sciences. Behav Res Methods 39, 175–191 (2007); published online EpubMay (10.3758/bf03193146).

