## Supplemental materials for "Intended Isocaloric Time-Restricted Eating Shifts Circadian Clocks but Does Not Improve Cardiometabolic Health in Women with Overweight"

Peters *et al.*

**The PDF file includes:**

Figs. S1 to S4

Tables S1 to S8

**Fig. S1. Individual Eating Times at the Baseline and during eTRE and lTRE.**

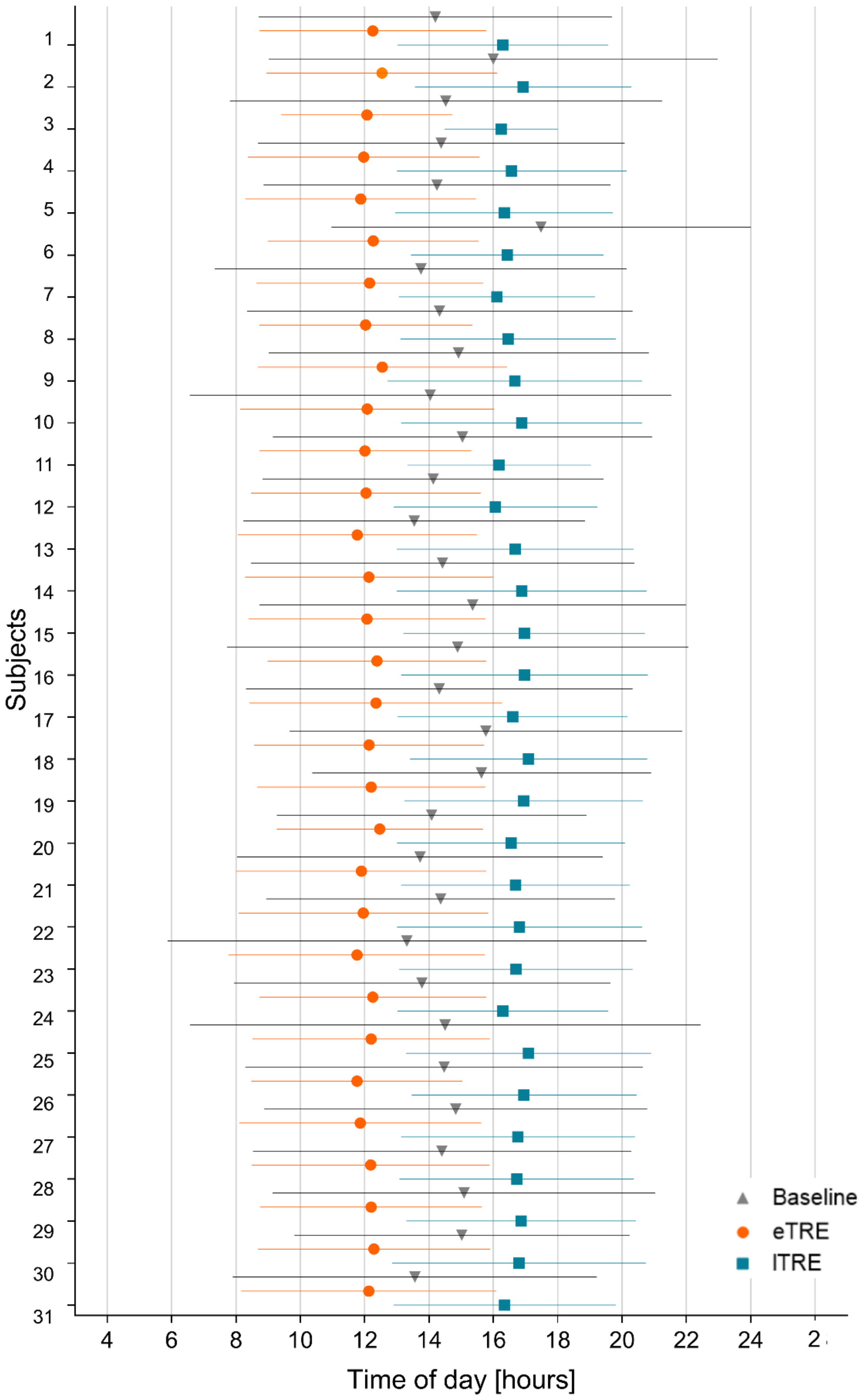


Plotted are individual times of day when participants started eating (left whisker) and stopped eating (right whisker) as well as the midpoint of eating (mean for each individual) in the baseline phase, eTRE and lTRE intervention phases.

**Fig. S2. Concentrations of Glucose, Hormones, and Free Fatty Acids in Oral Glucose Tolerance Test.**

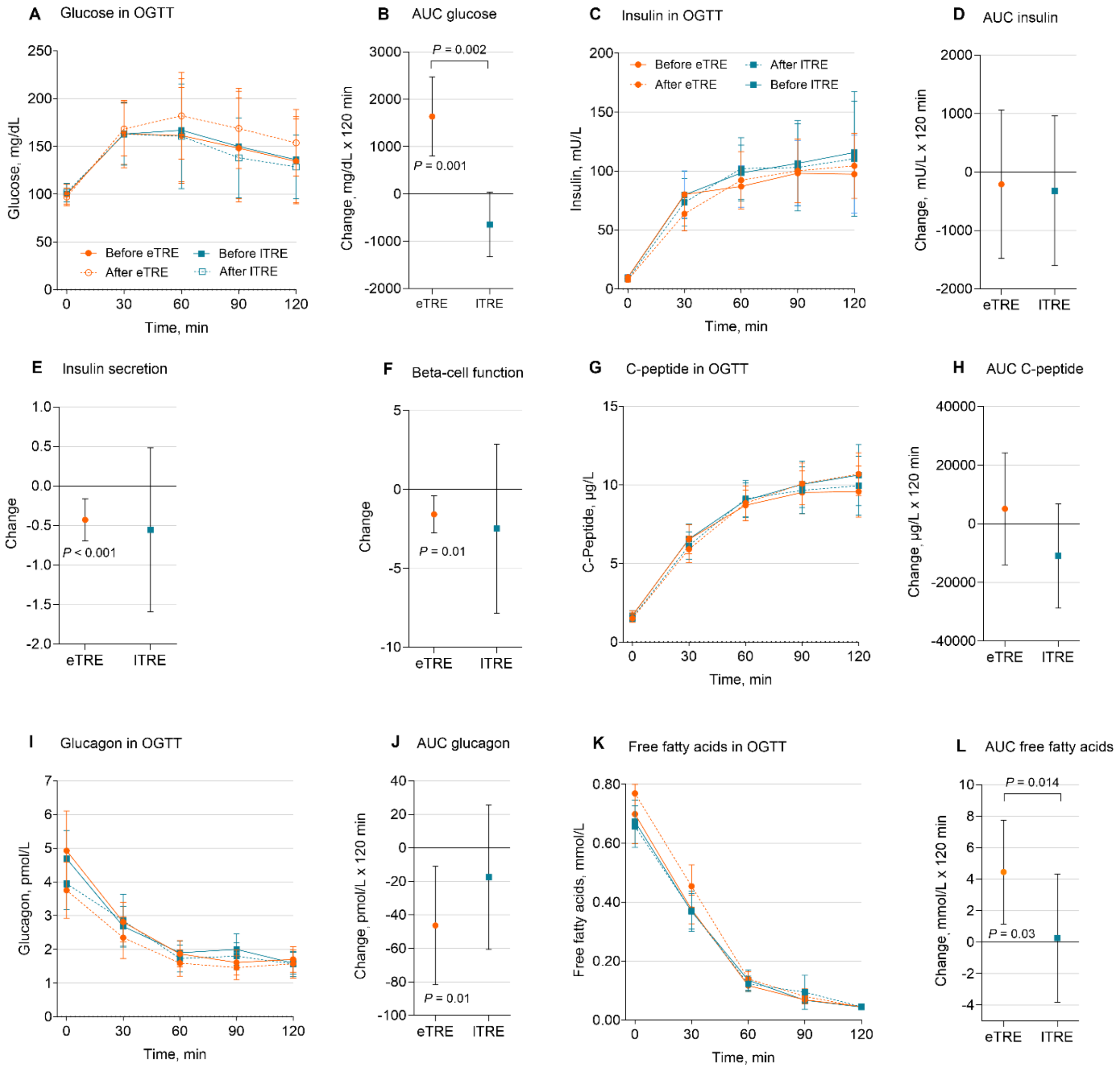


A, C, Presented are concentrations of glucose and insulin in the oral glucose tolerance test (OGTT) before and after eTRE and lTRE interventions (mean [SD]). B, D, Shown are changes in the areas under the curve (AUC) for glucose and insulin in OGTT within eTRE and lTRE as means with 95% CI. E, Changes of the insulin secretion (assessed by the insulinogenic index) are shown as means with 95% CI. F, Changes of the beta-cell function (assessed by the disposition index) are shown as means with 95% CI. G, I, K, Presented are concentrations of C-peptide, glucagon, and free fatty acids in the OGTT before and after eTRE and lTRE intervention (mean [SD]). H, J, L, Shown are changes in the AUCs for glucose, above mentioned hormones and fatty acids in OGTT within eTRE and lTRE as means with 95% CI.

**Fig. S3. Adipokine and Cytokine Concentrations.**

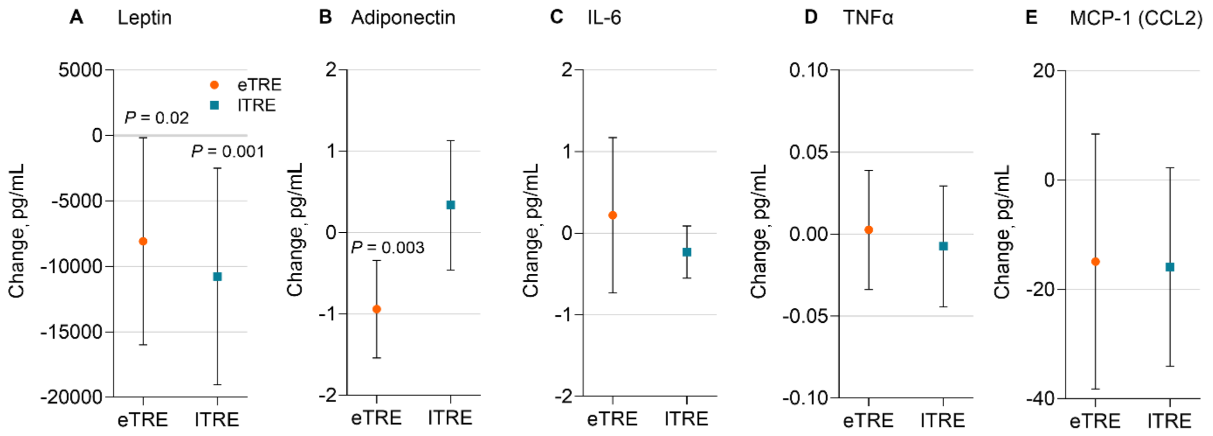


Presented are changes of adipokine and cytokine concentrations within eTRE and lTRE. A, leptin. B, adiponectin. C, IL-6. D, TNFα. E, MCP-1 (CCL2) (E). All data are shown as means with 95% CI.

**Fig. S4. Gene Expression in Peripheral Blood Mononuclear Cells.**

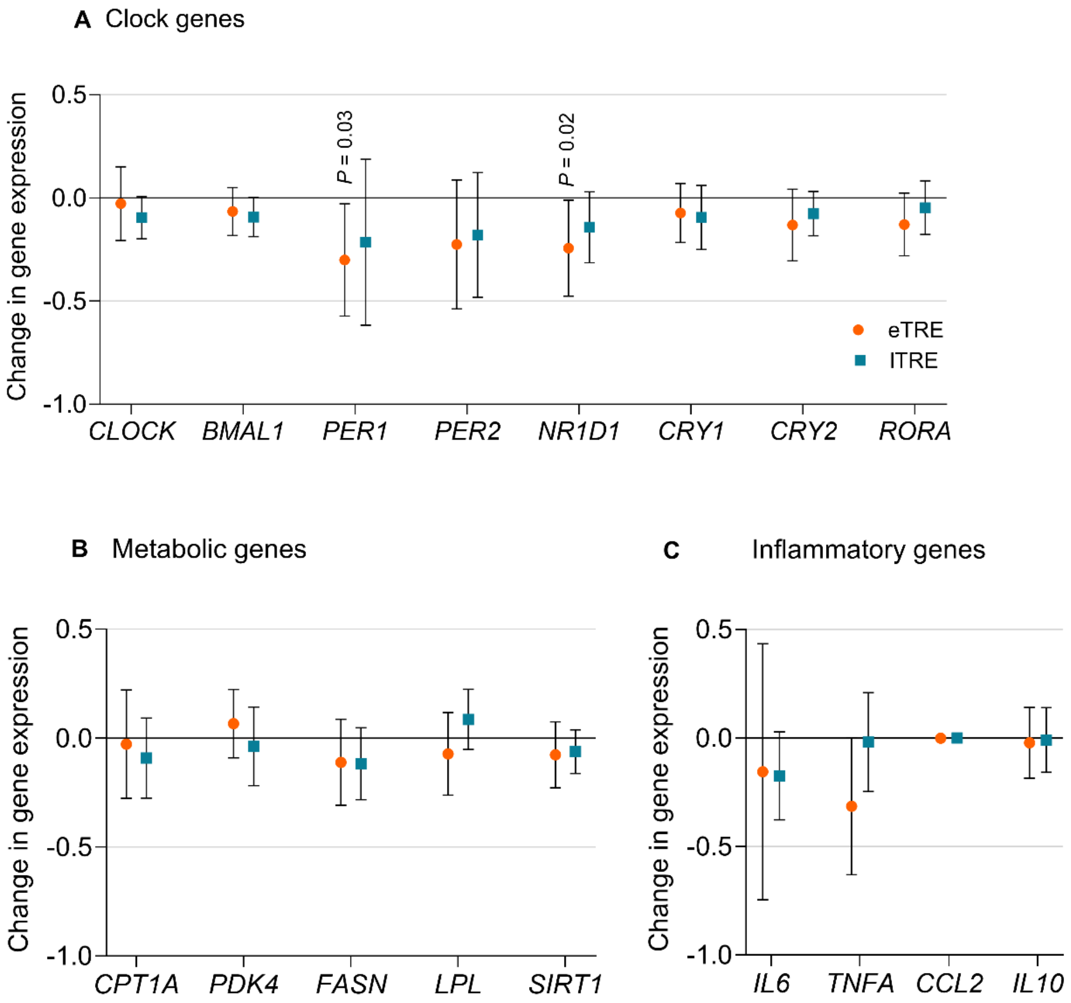


mRNA expression was measured by qPCR in peripheral blood mononuclear cells (PBMC) collected between 8:30 and 11:00 hr before and after each intervention. Changes in the expression of genes coding (A) clock genes, (B) key metabolic genes, and (C) inflammatory genes within eTRE and lTRE are shown as means with 95% CI.

Abbreviations: *CLOCK*, clock circadian regulator; *BMAL1*, basic helix-loop-helix ARNT like 1; *PER1*, period circadian regulator 1; *PER2*, period circadian regulator 2; *NR1D1*, nuclear receptor subfamily 1 group D member 1; *CRY1*, cryptochrome circadian regulator 1; *CRY2*, cryptochrome circadian regulator 2; *RORA*, RAR related orphan receptor A; *CPT1A*, carnitine palmitoyltransferase 1A; *PDK4*, pyruvate dehydrogenase kinase 4; *FASN*, fatty acid synthase; *LPL*, lipoprotein lipase; *SIRT1*, sirtuin 1; *IL6*, interleukin 6; *TNFA*, tumor necrosis factor alpha; *CCL2*, monocyte chemotactic protein 1; *IL10*, interleukin 10.

**Table S1. Adverse Events.**

| **Adverse events in eTRE (n = 10)** | **Adverse events in lTRE (n = 20)** |
|---|---|
| fatigue (n=1) | fatigue (n=3) |
| dizziness (n=1) | dizziness (n=3) |
| headaches (n=2) | headaches (n=3) |
| feeling cold (n=1) | feeling cold (n=1) |
| | sweeting in the evening (n=1) |
| vomiting after breakfast (n=1) | |
| | stomach pain (n=3) |
| | stomach grumbling (n=1) |
| | gastric reflux after the dinner (n=1) |
| night hypoglycemia (n=1) | |
| irregular defecation (n=3) | irregular defecation (n=3) |
| | diarrhea (n=1) |

The table shows self-reported adverse events possibly related to the intervention. No serious adverse events were reported.

**Table S2. Changes of Physical Activity.**

| | Change (95% CI) | | | | | |
|---|---|---|---|---|---|---|
| **Outcome** | **eTRE (n = 29)[a]** | **P value*** | **lTRE (n = 29)[a]** | **P value*** | **Difference between lTRE vs. eTRE interventions (95% CI)[a]** | **P value*** |
| MET | -0.02 (-0.05 to 0.01) | 0.19 | -0.01 (-0.04 to 0.02) | 0.52 | 0.01 (-0.02 to 0.04) | 0.54 |
| Light activity | 0.15 (-1.03 to 1.33) | 0.97 | -0.29 (-1.39 to 0.81) | 0.58 | -0.44 (-1.64 to 0.75) | 0.45 |
| Moderate activity | -0.51 (-1.34 to 0.32) | 0.22 | -0.26 (-1.26 to 0.74) | 0.59 | 0.25 (-0.74 to 1.23) | 0.61 |
| Sedentary activity | 0.35 (-1.29 to 2.00) | 0.16 | 0.56 (-0.85 to 1.96) | 0.21 | 0.20 (-1.42 to 1.83) | 0.97 |

Abbreviations: eTRE, early time-restricted eating; lTRE, late time-restricted eating; MET, metabolic equivalent of task

* Comparison by paired Student's t-test or Wilcoxon test

[a] Two baseline values for physical activity parameters were excluded in all pairwise comparisons due to two defect actigraphs at baseline.

**Table S3. Changes of Continuous Glucose Monitoring Indices.**

| | Change (95% CI) | | | | | |
|---|---|---|---|---|---|---|
| **Outcome** | **eTRE (n = 30)[a]** | **P value*** | **lTRE (n = 30)[a]** | **P value*** | **Difference between lTRE vs. eTRE interventions (95% CI)[a]** | **P value*** |
| MSG [mmol/L] | -0.03 (-0.26 to 0.21) | 0.82 | 0.12 (-0.08 to 0.31) | 0.23 | 0.14 (-0.06 tot 0.35) | 0.17 |
| SD [mmol/L] | 0.13 (0.02 to 0.24) | 0.003 | -0.03 (-0.09 to 0.03) | 0.34 | -0.16 (-0.28 to -0.05) | 0.002 |
| CV [%] | 2.78 (0.28 to 5.28) | 0.006 | -0.85 (-1.88 to 0.18) | 0.10 | -3.63 (-6.37 to -0.89) | 0.001 |
| MAGE [mmol/L] | 0.09 (-0.01 to 0.18) | 0.08 | -0.08 (-0.16 to 0.01) | 0.07 | -0.16 (-0.25 to -0.07) | 0.001 |
| CONGA [mmol/L] | 0.08 (-0.14 to 0.31) | 0.47 | 0.18 (0.00 to 0.36) | 0.05 | 0.10 (-0.08 to 0.29) | 0.27 |
| MODD [mmol/L] | -0.06 (-0.12 to 0.00) | 0.08 | -0.09 (-0.14 to -0.03) | 0.003 | -0.03 (-0.09 to 0.03) | 0.38 |
| LBGI | 2.84 (-2.29 to 7.97) | 0.18 | -0.44 (-1.02 to 0.13) | 0.24 | -3.28 (-8.56 to 1.99) | 0.04 |
| HBGI | 0.23 (0.08 to 0.37) | 0.006 | -0.02 (-0.17 to 0.12) | 0.40 | -0.25 (-0.43 to -0.08) | 0.003 |

Shown are changes of continuous glucose monitoring indices including mean sensor glucose, intra-day variation indices (SD, CV, MAGE, CONGA), inter-day variation indices (MODD) and risk scores (LBGI, HGBI) within and between-interventions.

Abbreviations: eTRE, early Time-Restricted Eating; lTRE, late Time-Restricted Eating; MSG, Mean Sensor Glucose; SD, Standard Deviation; CV, Percentage Coefficient of Variation for Glucose; MAGE, Mean Amplitude of Glycemic Excursions; CONGA, Continuous Overall Net Glycemic Action; MODD, Mean of Daily Differences; LBGI, Low Blood Glucose Index; HBGI, High Blood Glucose Index.

*Comparison by paired Student's t-test or Wilcoxon test

[a] One participant was excluded from CGM analysis due to incomplete CGM data. Data gaps can be attributed to non-regular measurement by the participant which avoided the saving of the data.

**Table S4. Changes of Oxidative Stress Markers.**

| Outcome | Change (95% CI) eTRE (n = 31) | P value* | lTRE (n = 31) | P value* | Difference between lTRE vs. eTRE interventions (95% CI) | P value** |
|---|---|---|---|---|---|---|
| Malondialdehyde, µmoL/L | 0.002 (-0.09 to 0.09) | 0.98 | 0.08 (-0.02 to 0.19) | 0.22 | 0.08 (-0.04 to 0.21) | 0.06 |
| 3-nitrotyrosine, pmol/mg | 0.03 (-1.32 to 1.39) | 0.95 | -0.08 (-1.03 to 0.86) | 0.73 | -0.12 (-1.64 to 1.40) | 0.17[a] |
| Protein carbonyls, nmol/mg | -0.05 (-0.13 to 0.03) | 0.23 | 0.02 (-0.06 to 0.11) | 0.60 | 0.07 (-0.06 to 0.20) | 0.26 |

Shown are changes of concentrations of oxidative stress markers malondialdehyde, 3-nitrotyrosine, and protein carbonyls within and between interventions.
Abbreviations: eTRE, early time-restricted eating; lTRE, late time-restricted eating
* Comparison by paired Student's t-test or Wilcoxon test
** Comparison of changes between eTRE and lTRE by the linear mixed model
[a] showed significant period effect

**Table S5. Feeling of Hunger and Satiety and Changes of Related Hormones.**

| Outcome | eTRE | P value* | lTRE | P value* | Difference between lTRE vs. eTRE interventions (95% CI) | P value* |
|---|---|---|---|---|---|---|
| **Hunger and Satiety Scores in the Morning** | | | | | | |
| Desire to eat, median (IQR)[a] | 4.40 (2.25-5.35) | | 1.20 (0.25-5.60) | | -1.49 (-2.58 to -0.40) | 0.02 |
| Hunger, median (IQR)[a] | 3.20 (2.10-4.80) | | 1.30 (0.15-4.30) | | -1.16 (-2.28 to -0.03) | 0.04 |
| Satiety, median (IQR)[a] | 5.10 (2.65-6.30) | | 5.40 (1.80-9.00) | | 0.90 (-0.45 to 2.24) | 0.22 |
| Capacity to eat, median (IQR)[a] | 5.00 (3.10-5.80) | | 2.90 (0.80-5.05) | | -1.16 (-2.28 to -0.03) | 0.03 |
| **Hunger and Satiety Scores in the Evening** | | | | | | |
| Desire to eat, median (IQR)[a] | 4.50 (2.20-7.65) | | 3.30 (0.70-5.75) | | -0.85 (-1.82 to 0.12) | 0.10 |
| Hunger, median (IQR)[a] | 3.60 (1.95-6.55) | | 3.20 (0.25-5.15) | | -0.58 (-1.68 to 0.53) | 0.25 |
| Satiety, median (IQR)[a] | 5.00 (2.75-7.85) | | 6.70 (3.35-9.00) | | 0.75 (-0.52 to 2.08) | 0.12 |
| Capacity to eat, median (IQR)[a] | 4.40 (2.60-5.40) | | 3.30 (1.00-5.35) | | -0.55 (-1.59 to 0.48) | 0.28 |
| **Changes of Hormones Related to Hunger and Satiety** | | | | | | |
| Peptide YY[b], 95% CI | -13.0 (-24.5 to -1.5) | 0.01 | 22.5 (13.1 to 31.9) | $1.6\times10^{-4}$ | 34.7 (18.2 to 51.3) | $1.2\times10^{-4}$** |
| Ghrelin[c], 95% CI | 1.31 (-2.30 to 4.92) | 0.50 | -0.10 (-1.91 to 1.72) | 0.59 | -1.36 (-5.42 to 2.69) | 0.84** |

Shown are absolute hunger and satiety scores assessed by a visual analog scale in the morning and in the evening in eTRE and lTRE and changes of hormones related to hunger and satiety (PYY, ghrelin) within and between interventions.

Abbreviations: eTRE, early time-restricted eating; lTRE, late time-restricted eating.

* Comparison by paired Student's t-test or Wilcoxon test

** Comparison of changes between eTRE and lTRE by the linear mixed model

[a] Two participants in each intervention were identified were excluded for paired analysis (eTRE n = 29; lTRE n = 29; Difference between TRE interventions n = 27).

[b] One participant in each intervention were identified as outliner and therefore were excluded for paired analysis (eTRE n = 30; lTRE n = 30; Difference between TRE interventions n = 29).

[c] Two participants in both interventions and one further participant in lTRE intervention were identified as outliner and therefore were excluded for paired analysis (eTRE n = 29; lTRE n = 28; Difference between TRE interventions n = 28)

**Table S6. Baseline Characteristics in Subjects with Impaired Glucose Metabolism.**

| | Subjects with NGT (n = 18) | Subjects with IFG/IGT (n =13) |
|---|---|---|
| Age, mean (SD), y | 55 (9) | 63 (5) |
| Sex, female:male | 18:0 | 11:0 |
| Race, white:other | 18:0 | 11:0 |
| Ethnicity: Caucasian:other | 18:0:0 | 11:0:0 |
| Weight, mean (SD), kg | 80.9 (7.6) | 84.8 (9.3) |
| Fat mass, mean (SD), kg | 31.7 (6.2) | 37.4 (5.9) |
| Lean mass, mean (SD), kg | 49.1 (5.9) | 47.4 (4.7) |
| Waist circumference, mean (SD), cm | 96 (8) | 104 (8) |
| BMI, mean (SD), $kg/m^2$ | 29.2 (2.6) | 32.2 (2.4) |
| **Glucose homeostasis** | | |
| Fasting glucose, mean (SD), mg/dL | 87 (7) | 101 (12) |
| Glucose 120 min in OGTT, mean (SD), mg/dL | 101 (17) | 153 (29) |
| HbA1c, mean (SD), %[a] | 5.35 (0.26) | 5.64 (0.33) |
| **Cardiometabolic parameters** | | |
| Systolic blood pressure, median (IQR), mm Hg | 114 (110-125) | 120 (115-139) |
| Diastolic blood pressure, median (IQR), mm Hg | 76 (69-78) | 73 (68-80) |
| Total cholesterol, mean (SD), mmol/L | 5.75 (0.98) | 5.47 (0.84) |
| HDL cholesterol, mean (SD), mmol/L | 1.64 (0.43) | 1.42 (0.27) |
| LDL cholesterol, mean (SD), mmol/L | 3.52 (0.82) | 3.41 (0.86) |
| Triglycerides, mean (SD), mmol/L | 1.30 (0.77) | 1.42 (0.35) |
| ASAT, mean (SD), U/L | 30.1 (6.0) | 25.2 (5.9) |
| ALAT, median (IQR), U/L | 21.8 (17.2-23.8) | 26.6 (21.0-29.0) |
| GGT, median (IQR), U/L | 19.4 (15.8-24.3) | 24.1 (20.4-31.9) |
| hsCRP, median (IQR), mg/L | 1.15 (0.50-3.05) | 1.60 (1.30-2.45) |

Abbreviations: eTRE, early time-restricted eating; lTRE, late time-restricted eating; WHR, waist-to-hip ratio; BMI, body mass index; NGT, normal glucose tolerance; IFG, impaired fasting glucose; IGT, impaired glucose tolerance; HbA1c, hemoglobin $A_{1C}$, HDL, high-density lipoprotein; hsCRP, high-sensitive C-reactive protein; LDL, low-density lipoprotein.
SI conversion factors: To convert cholesterol mmol/L to mg/dL divide by 0.0259; glucose mg/dL to mmol/L multiply by 0.0555; triglycerides mg/dL to mmol/L multiply by 0.0113.
[a] One baseline value in group B (lTRE-eTRE) was not measured due to loss of the sample.

**Table S7. TRE Effects on Body Composition, Glucose Homeostasis, and Cardiometabolic Parameters in Subjects with Impaired Glucose Metabolism.**

| Outcome | Change (95% CI) | | | | Difference between lTRE vs. eTRE interventions (95% CI) | P value** |
|---|---|---|---|---|---|---|
| | eTRE (n = 13) | P value* | lTRE (n = 13) | P value* | | |
| **Body Composition** | | | | | | |
| Weight, kg | -1.45 (-1.96 to -0.93) | 0.002 | -0.73 (-1.24 to -0.22) | 0.005 | 0.72 (-0.06 to 1.49) | 0.04 |
| Fat mass, kg[a] | -0.82 (-1.78 to 0.14) | 0.08 | 0.16 (-0.44 to 0.76) | 0.56 | 0.98 (0.12 to 1.84) | 0.31 |
| Lean mass, kg[a] | -0.76 (-1.94 to -0.42) | 0.05 | -0.48 (-1.13 to 0.17) | 0.09 | 0.28 (-0.68 to 1.24) | 0.22 |
| Waist circumference, cm | -1.05 (-2.78 to 0.67) | 0.21 | -0.87 (-2.88 to 1.15) | 0.36 | 0.19 (-2.32 to 2.69) | 0.34 |
| BMI, $kg/m^2$ | -0.63 (-0.78 to -0.48) | $9.7 \times 10^{-7}$ | -0.16 (-0.28 to -0.06) | 0.007 | 0.46 (0.29 to 0.64) | $4.7 \times 10^{-4}$ |
| **Glucose Homeostasis** | | | | | | |
| Insulin sensitivity[b] | 0.29 (-0.33 to 0.91) | 0.28 | 0.21 (-0.21 to 0.64) | 0.35 | -0.08 (-0.95 to 0.79) | 0.30 |
| Insulin secretion[c] | -0.35 (-0.70 to -0.01) | 0.007 | 0.03 (-0.13 to 0.20) | 0.42 | 0.39 (-0.01 to 0.79) | 0.38 |
| Beta-cell function[d] | -1.03 (-2.89 to 0.83) | 0.15 | 0.55 (-0.08 to 1.17) | 0.10 | 1.58 (-0.60 to 3.77) | 0.62 |
| Fasting glucose, mg/dL | -0.05 (-0.36 to 0.25) | 0.72 | 0.04 (-0.15 to 0.23) | 0.46 | 0.09 (-0.24 to 0.42) | 0.29 |
| AUC glucose, mg/dL x 120 min | 865 (-413 to 2144) | 0.17 | -1213 (-2380 to -45) | 0.04 | -2078 (-3768 to 388) | 0.22 |
| Fasting insulin, mU/L | -1.91 (-3.78 to -0.03) | 0.02 | -2.77 (-8.15 to 2.60) | 0.20 | -0.87 (-6.71 to 4.98) | 0.74 |
| AUC insulin, mU/L x 120 min | -1372 (-3828 to 1084) | 0.51 | 368 (-2491 to 3227) | 0.75 | 1740 (-2077 to 5557) | 0.22 |
| $HbA_{1c}$, % | -0.01 (-0.08 to 0.06) | 0.77 | -0.03 (-0.08 to 0.03) | 0.30 | 0.02 (-0.07 to 0.10) | 0.74 |
| MSG, mmol/L | -0.12 (-0.42 to 0.18) | 0.40 | -0.05 (-0.34 to 0.23) | 0.70 | 0.07 (-0.20 to 0.34) | 0.59* |
| **Cardiometabolic Parameters** | | | | | | |
| Systolic blood pressure, mm Hg | 1.80 (-4.61 to 8.21) | 0.54 | 6.54 (-1.64 to 14.72) | 0.11 | 2.20 (-9.86 to 14.26) | 0.96 |

| | | | | | | |
|---|---|---|---|---|---|---|
| Diastolic blood pressure, mm Hg | 0.40 (-6.52 to 7.32) | 0.90 | 0.62 (-5.15 to 6.38) | 0.82 | -2.00 (-12.09 to 8.09) | 0.35 |
| Total cholesterol, mmol/L | -0.22 (-0.57 to 0.13) | 0.19 | -0.12 (-0.29 to 0.05) | 0.16 | 0.11 (-0.33 to 0.54) | 0.58 |
| HDL cholesterol, mmol/L | -0.09 (-0.14 to -0.04) | 0.003 | -0.09 (-0.15 to -0.04) | 0.008 | 0.002 (-0.05 to 0.05) | 0.30 |
| LDL cholesterol, mmol/L | -0.72 (-0.50 to 0.35) | 0.72 | 0.19 (-0.16 to 0.54) | 0.27 | 0.26 (-0.44 to 0.96) | 0.74 |
| Triglycerides, mmol/L | 0.06 (-0.14 to 0.26) | 0.51 | -0.10 (-0.31 to 0.11) | 0.32 | -0.16 (-0.42 to 0.09) | 0.30 |
| ASAT, U/L | -0.50 (-0.82 to 1.82) | 0.42 | -1.39 (-3.36 to 0.58) | 0.15 | -1.89 (-4.41 to 0.62) | 0.07 |
| ALAT, U/L | 0.42 (-1.02 to 1.86) | 0.53 | -3.52 (-10.06 to 3.02) | 0.20 | -3.95 (-11.47 to 3.58) | 0.16 |
| GGT, U/L | -3.21 (-5.62 to -0.80) | 0.008 | -3.05 (-6.40 to 0.30) | 0.06 | 0.15 (-4.33 to 4.64) | 0.89 |
| **Adipokines, Inflammatory Markers, and Oxidative Stress Markers** | | | | | | |
| Leptin, pg/mL | -14954 (-31015 to 1106) | 0.046 | -19537 (-33645 to -5429) | 0.002 | -4583 (-20391 to 11226) | 0.31 |
| Adiponectin, pg/mL | -0.001 (-1.20 to 1.20) | 0.10 | -0.91 (-2.02 to 0.19) | 0.99 | -0.91 (-2.57 to 0.75) | 0.89 |
| IL-6, pg/mL | -0.23 (-0.82 to 0.37) | 0.44 | 0.83 (-1.55 to 3.22) | 0.92 | -1.06 (-3.46 to 1.33) | 0.99 |
| TNFα, pg/mL | -0.003 (-0.04 to 0.04) | 0.75 | -0.01 (-0.06 to 0.04) | 0.69 | -0.007 (-0.06 to 0.05) | 0.45 |
| MCP-1, pg/mL | -20.10 (-64.39 to 24.19) | 0.32 | -27.61 (-61.99 to 6.77) | 0.11 | -7.51 (-40.74 to 55.77) | 0.78 |
| hsCRP, mg/L | 0.08 (-2.02 to 2.18) | 0.59 | -0.07 (-0.38 to 0.24) | 0.61 | -0.15 (-2.29 to 1.98) | 0.16 |
| Malondialdehyde, µmoL/L | -0.06 (-0.24 to 0.12) | 0.63 | 0.01 (-0.11 to 0.13) | 0.81 | -0.07 (-0.28 to 0.14) | 0.23 |
| 3-nitrotyrosine, pmol/mg | -2.67 (-4.60 to -0.73) | 0.02 | -0.15 (-1.34 to 1.05) | 0.77 | 2.52 (0.60 to 4.44) | 0.10[e] |
| Protein carbonyls, nmol/mg | -0.02 (-0.12 to 0.09) | 0.76 | 0.10 (-0.04 to 0.23) | 0.14 | -0.11 (-0.27 to 0.05) | 0.32 |

Abbreviations: WHR, waist-to-hip ratio; BMI, body mass index; HDL, high-density lipoprotein; hsCRP, high sensitive C-reactive protein; LDL, low-density lipoprotein; ASAT, aspartate-aminotransferases; ALAT, alanine-aminotransferases; GGT, gamma-glutamyltransferases; MSG, mean sensor glucose

* Comparison by paired Student's t-test or Wilcoxon test

** Comparison of changes between eTRE and lTRE by the linear mixed model

[a] one eTRE and two lTRE data sets (pre and post) were excluded because of invalid bioelectrical impedance analysis

[b] assessed by Matsuda index in OGTT
[c] assessed by insulinogenic index in OGTT
[d] assessed by disposition index in OGTT
[e] showed significant carryover effect, therefore only the data from the first intervention included in analysis

### Table S8. Primer Sequences.

| Gene name | Gene symbol | Forward primer | Reverse primer |
|---|---|---|---|
| **Clock genes** | | | |
| Clock circadian regulator | *CLOCK* | ATTCCACAAGGCATGTCCCA | TTTGCTTCTATCATGCGTGTCC |
| Basic helix-loop-helix ARNT like 1 | *BMAL1* | CATTAAGAGGTGCCACCAATCC | CAAAAATCCATCTGCTGCCC |
| Period circadian regulator 1 | *PER1* | ATTCCGCCTAACCCCGTATGT | CCGCGTAGTGAAAATCCTCTTG |
| Period circadian regulator 2 | *PER2* | AGCAGGTGAAAGCCAATGAAG | AGGTAACGCTCTCCATCTCCTC |
| Nuclear receptor subfamily 1 group D member 1 | *NR1D1* | TGACCTTTCTCAGCATGACCAA | CAAAGCGCACCATCAGCAC |
| Cryptochrome circadian regulator 1 | *CRY1* | GGGACCTGTGGATTAGTTGGG | GCTCCAATCTGCATCAAGCAA |
| Cryptochrome circadian regulator 2 | *CRY2* | TGCATCTGTTGACACTCATGATTC | GGTACTCCCCCAGCCCAG |
| RAR related orphan receptor A | *RORA* | ACTCCTGTCCTCGTCAGAAGA | CATCCCTACGGCAAGGCATTT |
| **Metabolic genes** | | | |
| Carnitine palmitoyltransferase 1A | *CPT1A* | ATTATGCCATGGATCTGCTG | AGCGGAGCAGAGTGGAATC |
| Fatty acid synthase | *FASN* | AGACACTCGTGGGCTACAGCAT | ATGGCCTGGTAGGCGTTCT |
| Pyruvate dehydrogenase kinase 4 | *PDK4* | CCCTGAGAATTATTGACCGCCT | AAGCCGTAACCAAAACCAGCC |
| Sirtuin 1 | *SIRT1* | ATGCTGGCCTAATAGAGTGGCA | CCTCAGCGCCATGGAAAAT |
| Lipoprotein lipase | *LPL* | TGCAGGAAGTCTGACCAATAAG | CCCTCTGGTGAATGTGTGTAAG |
| **Inflammation genes** | | | |
| Interleukin 6 | *IL6* | AGCCCTGAGAAAGGAGACATGTA | TCTGCCAGTGCCTCTTTGCT |
| Interleukin 10 | *IL10* | ACGGCGCTGTCATCGATT | GGCATTCTTCACCTGCTCCA |
| Tumor necrosis factor alpha | *TNFα* | GGACCTCTCTCTAATCAGCCCTC | TCGAGAAGATGATCTGACTGCC |
| Monocyte chemotactic protein 1 | *CCL2* | CATAGCAGCCACCTTCATTCC | TCTGCACTGAGATCTTCCTATTGG |
| **Housekeeper gene** | | | |
| Beta-2-microglobulin | *B2M* | CTATCCAGCGTACTCCAAAG | AAACCCAGACACATAGCAAT |

Sequences of primers used for the quantitative real-time PCR in PBMC are shown. Genes were selected based on the previously published human or animal data on their circadian rhythmicity and/or their potential role in the TRE-induced metabolic effects.
